# High-dimensional profiling reveals phenotypic heterogeneity and disease-specific alterations of granulocytes in COVID-19

**DOI:** 10.1101/2021.01.27.21250591

**Authors:** Magda Lourda, Majda Dzidic, Laura Hertwig, Helena Bergsten, Laura M. Palma Medina, Egle Kvedaraite, Puran Chen, Jagadeeswara R. Muvva, Jean-Baptiste Gorin, Martin Cornillet, Johanna Emgård, Kirsten Moll, Marina García, Kimia T. Maleki, Jonas Klingström, Jakob Michaëlsson, Malin Flodström-Tullberg, Susanna Brighenti, Marcus Buggert, Jenny Mjösberg, Karl-Johan Malmberg, Johan K. Sandberg, Jan-Inge Henter, Elin Folkesson, Sara Gredmark-Russ, Anders Sönnerborg, Lars I. Eriksson, Olav Rooyackers, Soo Aleman, Kristoffer Strålin, Hans-Gustaf Ljunggren, Niklas K. Björkström, Mattias Svensson, Andrea Ponzetta, Anna Norrby-Teglund, Benedict J. Chambers, the Karolinska KI/K COVID-19 Study Group

## Abstract

Since the outset of the COVID-19 pandemic, increasing evidence suggests that the innate immune responses play an important role in the disease development. A dysregulated inflammatory state has been proposed as key driver of clinical complications in COVID-19, with a potential detrimental role of granulocytes. However, a comprehensive phenotypic description of circulating granulocytes in SARS-CoV-2-infected patients is lacking. In this study, we used high-dimensional flow cytometry for granulocyte immunophenotyping in peripheral blood collected from COVID-19 patients during acute and convalescent phases. Severe COVID-19 was associated with increased levels of both mature and immature neutrophils, and decreased counts of eosinophils and basophils. Distinct immunotypes were evident in COVID-19 patients, with altered expression of several receptors involved in activation, adhesion and migration of granulocytes (e.g. CD62L, CD11a/b, CD69, CD63, CXCR4). Paired sampling revealed recovery and phenotypic restoration of the granulocytic signature in the convalescent phase. The identified granulocyte immunotypes correlated with distinct sets of soluble inflammatory markers supporting pathophysiologic relevance. Furthermore, clinical features, including multi-organ dysfunction and respiratory function, could be predicted using combined laboratory measurements and immunophenotyping. This study provides a comprehensive granulocyte characterization in COVID-19 and reveals specific immunotypes with potential predictive value for key clinical features associated with COVID-19.

**Significance:** Accumulating evidence shows that granulocytes are key modulators of the immune response to SARS-CoV-2 infection and their dysregulation could significantly impact COVID-19 severity and patient recovery after virus clearance. In the present study, we identify selected immune traits in neutrophil, eosinophil and basophil subsets associated to severity of COVID-19 and to peripheral protein profiles. Moreover, computational modeling indicates that the combined use of phenotypic data and laboratory measurements can effectively predict key clinical outcomes in COVID-19 patients. Finally, patient-matched longitudinal analysis shows phenotypic normalization of granulocyte subsets 4 months after hospitalization. Overall, in this work we extend the current understanding of the distinct contribution of granulocyte subsets to COVID-19 pathogenesis.

## Introduction

Caused by the novel coronavirus SARS-CoV-2, the current COVID-19 pandemic poses an unprecedented threat to the global health systems, with over 2 million confirmed deaths reported worldwide in January 2021 (https://www.who.int/emergencies/diseases/novel-coronavirus-2019). About 80% of COVID-19 patients present with a mild or moderate disease course, while 15-20% develop severe complications (1, 2), including respiratory failure, coagulation abnormalities and life-threatening acute respiratory distress syndrome (ARDS). A dysregulated inflammatory state has been proposed as a key driver of clinical complications of COVID-19 (3) and has been associated with increased mortality (4). High levels of several pro-inflammatory mediators (e.g. Interleukin-1β (IL-1β), IL-6, Tumor necrosis factor α(TNF α) and CXCL8) are detected early after viral infection (5), and the resolution of this inflammatory response seems impaired in patients with severe disease progression (5, 6). Many of these pro-inflammatory cytokines are associated with granulocyte activation and recruitment (7), and it has been reported that an increased neutrophil-to-lymphocyte ratio (NLR) in peripheral blood can be used as a prognostic marker of higher disease severity in COVID-19 patients, similar to SARS-CoV-1 and MERS (8-10).

The cellular components of the polymorphonuclear granulocyte family – neutrophils, eosinophils and basophils – arise from a common myeloid progenitor in humans (11) and represent the very first line of defense against invading microbes. The role of granulocytes in viral infection has mostly been confined to neutrophils (12, 13), though eosinophils and basophils have been implicated in the host response to viruses as well (14, 15). In COVID-19, an increased neutrophil abundance in circulation is mirrored by a substantial enrichment of granulocytes in the inflamed lung (16, 17). Several reports highlighted the presence of immature neutrophils in blood of COVID-19 patients (18, 19), implying a marked emergency granulopoiesis occurring in the bone marrow. The presence of immature neutrophil populations is phenotypically reminiscent of granulocytic myeloid-derived suppressor cells (G-MDSCs), and has thus been linked to a potential immunosuppressive role of neutrophils during SARS-CoV-2 infection (19, 20). A confounding factor in many of the reported studies is represented by the broad use of peripheral blood mononuclear cells (PBMC) (e.g. (6, 18, 21)), where only the low-density granulocytic fraction can be captured, which could lead to an inherent bias in the result interpretation (22, 23). Longitudinal studies performed during the course of SARS-CoV-2 infection have revealed that neutrophil numbers slowly decrease during infection, and the rate of such decrease is lower in more severe patients (6). Interestingly, basophil and eosinophil counts were found to be inversely correlated with neutrophil levels (24).

In the present study we set out to delineate the phenotypic alterations within neutrophil, eosinophil and basophil populations during the early phase of SARS-CoV-2 infection in patients with acute COVID-19 and in the convalescent phase. We provide a detailed analysis at the single-cell resolution of granulocyte diversity in fresh whole blood of COVID-19 patients using 25-color flow cytometry, integrated with soluble factor detection by proteomics and detailed clinical information. Overall, this study delineates the biological contribution of granulocyte subsets to COVID-19 pathogenesis, with an emphasis on their phenotypical heterogeneity in viral infection. The data reveals specific immunotypes with potential predictive value for key clinical features associated with COVID-19 severity.

## Results

### Altered composition of granulocyte populations is associated with disease severity in COVID-19

To provide an in-depth characterization of the granulocyte compartment in patients with moderate or severe COVID-19, we designed a strategy based on the integrated analysis of high dimensional flow cytometry data from whole blood cells, extensive proteomic screening in serum and plasma, and detailed clinical and laboratory information from patients included in the study (Figure 1A). Twenty-six hospitalized COVID-19 patients (10 moderate and 16 severe) and age-matched healthy controls were recruited at Karolinska University Hospital as part of the Karolinska KI/K COVID-19 Immune Atlas effort (25-28) (Figure 1B, Supplemental Tables 1-3). No differences in age, sex, body-mass index (BMI) or time from symptom debut until hospital admission, were present between moderate and severe patients (Figure 1B, Supplemental Table 2). The severe group displayed higher concentration of the inflammatory markers D-dimer and C-reactive protein (CRP), coagulation and inflammatory markers, and included all patients (n=4) who subsequently died in the hospital (Figure 1B).

**Figure 1.**
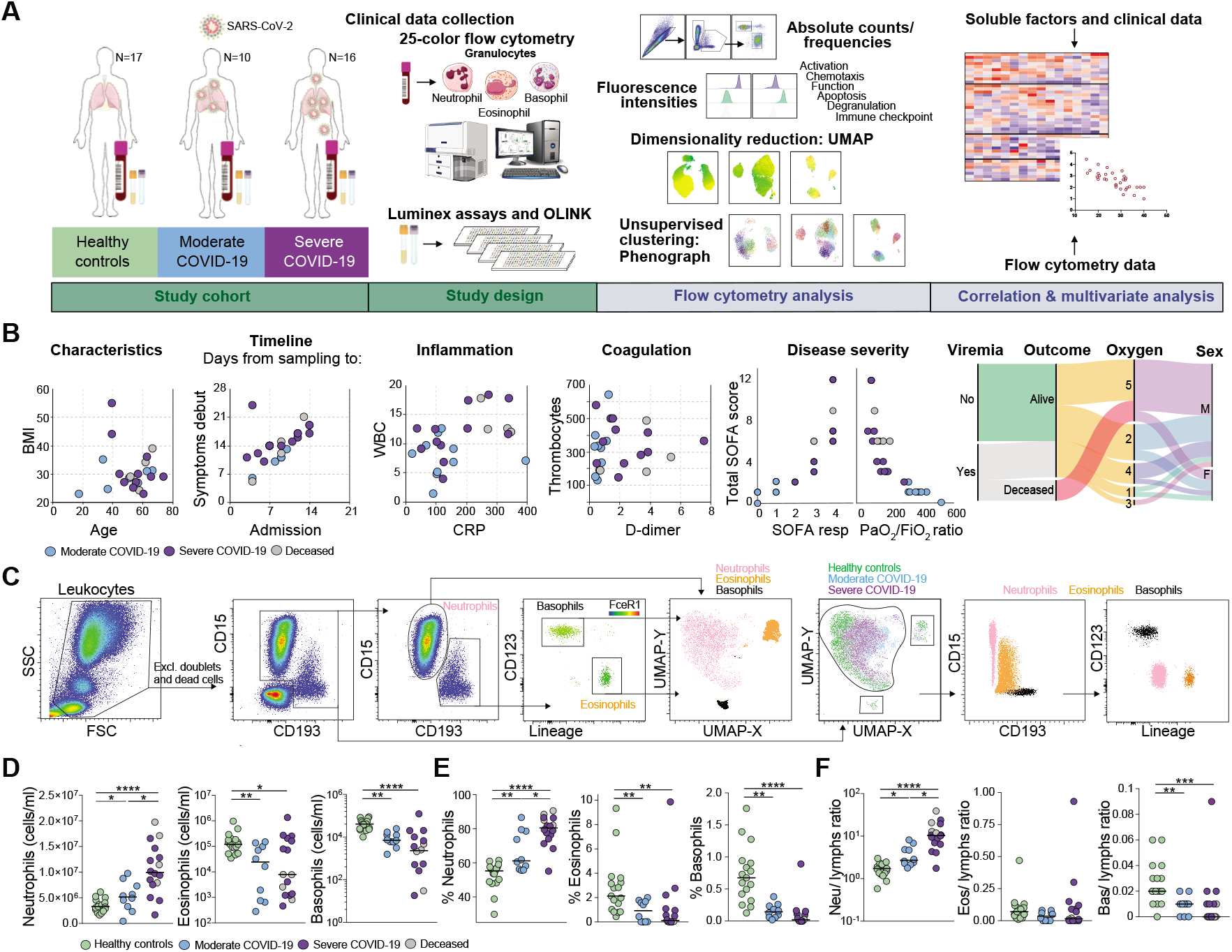
Disease severity-dependent neutrophilia and marked decrease of eosinophils and basophils in COVID-19. (**A**) Experimental and analytical workflow of the study performed on samples from moderate and severe COVID-19 patients and age-matched healthy controls. (**B**) Dot plots (left) and alluvial diagram (right) describing demographics and the clinical characteristics of the patients included in the study cohort. Severity groups (moderate=blue; severe=purple; deceased=gray) are indicated. (**C**) Gating strategy for the identification of granulocyte subsets and their UMAP projection is shown on the left. On the right, analogue results are obtained through unsupervised subset identification based on UMAP projection of total granulocytes. (**D-E**) Absolute cell counts (**D**) and frequencies (**E**) among total leukocytes for neutrophils, eosinophils and basophils in healthy controls (n=17), moderate COVID-19 patients (n=10) and severe COVID-19 patients (n=16). (**F**) Ratios of granulocyte absolute counts over lymphocyte absolute counts in healthy controls (n=17), moderate COVID-19 patients (n=10) and severe COVID-19 patients (n=16). In (**D–F**), Kruskall-Wallis test and two-stage Benjamini, Krieger and Yekutieli test. Bars represent median. * p < 0.05; ** p < 0.01; *** p < 0.001; **** p < 0.0001. Neu, neutrophils; eos, eosinophils; bas, basophils; lymphs, lymphocytes.

A 25-color flow cytometry panel was applied on freshly isolated whole blood cells from COVID-19 patients and healthy controls (Figure 1C) to identify and characterize the main granulocyte subsets. To minimize batch-effects, samples were processed and acquired during a short period of time (3 weeks). A manual gating strategy for neutrophils, eosinophils and basophils was used based on expression of canonical lineage-defining markers (Figure 1C). Neutrophils were identified as CD15^+^CD193^-^CD66b^+^CD16^bright/dim^, eosinophils as CD15^bright/dim^CD193^bright/dim^CD16^-^FceR1^dim^CD123^dim/-^ and basophils as CD15^-^ CD193^bright^FceR1^bright^CD123^bright^HLA-DR^dim/-^. Projection of gated cells into the Uniform Manifold Approximation and Projection (UMAP) space integrating all the analyzed markers confirmed the efficacy of the manual gating strategy (Figure 1C).

The analysis of absolute leukocyte counts in COVID-19 patients highlighted relevant alterations in the granulocyte compartment in peripheral blood. In particular, neutrophils were significantly increased in patients with severe disease course, while eosinophils and basophils were decreased in COVID-19 patients when compared to non-infected healthy controls (Figure 1D). A similar trend was observed in the relative frequencies of granulocyte subsets over total leukocytes. A higher proportion of neutrophils in patients with moderate and severe COVID-19 (66.65 ± 11.71% and 78.96 ± 8.83 respectively) was associated with the reduction of the eosinophil and basophil fractions (up to 3 and 7.5-fold, respectively, Figure 1E). The ratios of neutrophils over eosinophils or basophils increased in both patient groups, while no difference in the eosinophil to basophil ratios was observed between controls and patients (Supplemental Figure 1A). In line with previous reports (29), we found that severe COVID-19 was associated with higher neutrophil-to-lymphocyte and neutrophil-to-T cell ratios, as determined by absolute cell count (Figure 1F, Supplemental Figure 1B). We observed an opposite trend for basophils, which was associated with disease severity, while we did not detect any significant alteration in the eosinophil to lymphocyte or eosinophil to T cell ratios (Figure 1F, Supplemental Figure 1B).

This set of data highlights that the granulocyte expansion observed during acute SARS-CoV-2 infection is dominated by neutrophils and is coupled to the reduction of the physiological levels of circulating eosinophils and basophils.

### Severe COVID-19 is characterized by the emergence of immature neutrophils in peripheral blood

Neutrophils have been shown to display a high degree of phenotypical heterogeneity, which has important implications on their function in several pathological contexts (30). Different neutrophil maturation states are associated with major phenotypic differences, e.g. CD16 upregulation during development (23, 30). In COVID-19 patients, particularly in those with severe disease, we observed a robust enrichment of a neutrophil subset characterized by low expression of CD16 suggesting that these were immature CD16^dim^ neutrophils (Figure 2A).

**Figure 2.**
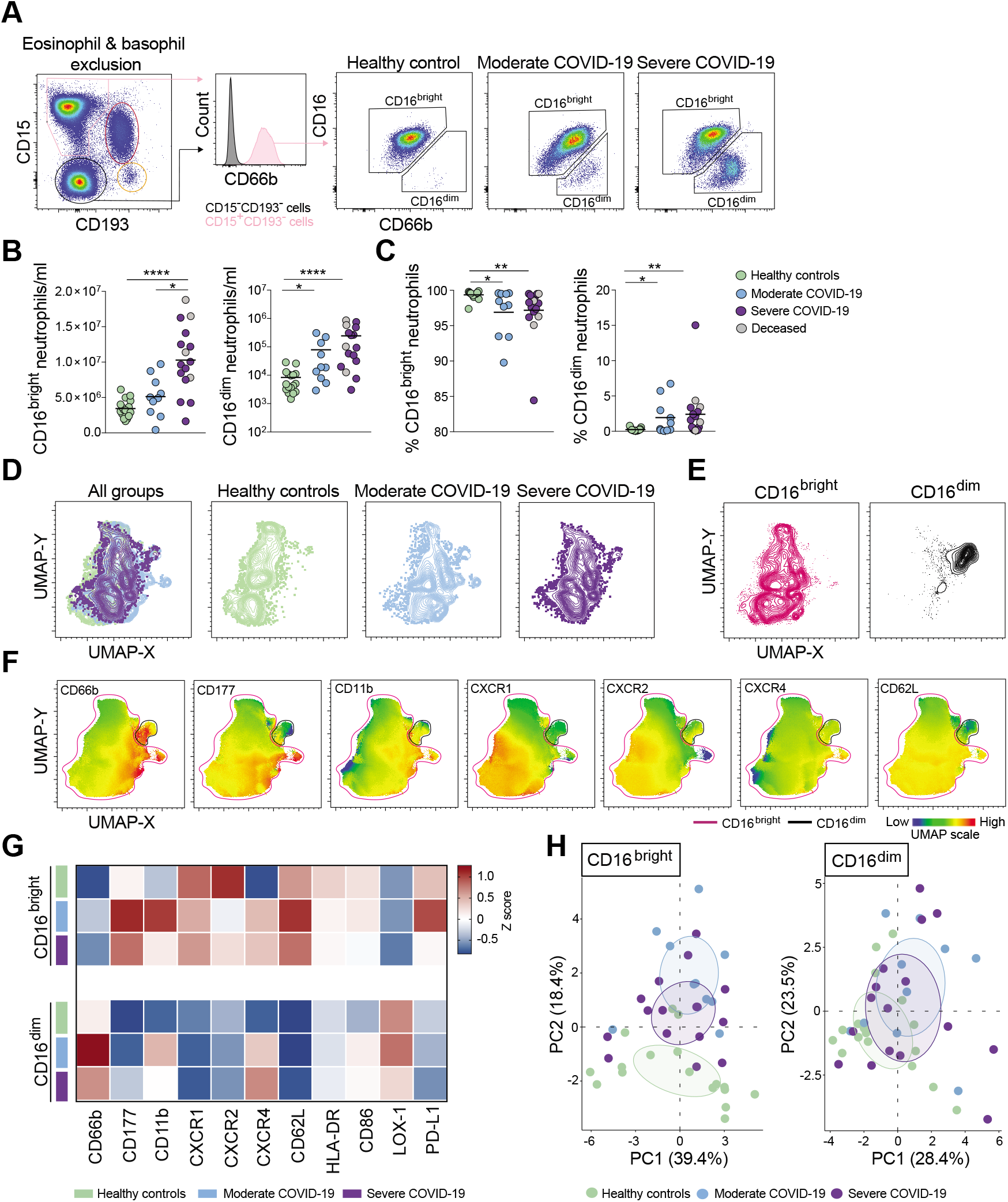
Emergence of immature neutrophils in severe COVID-19. (**A**) Representative plots showing neutrophil identification and CD16^bright^ and CD16^dim^ subset discrimination in different study groups. Absolute cell counts (**B**) and frequencies (**C**) among total neutrophils of CD16^bright^ and CD16^dim^ cells in healthy controls (n=17), moderate COVID-19 patients (n=10) and severe COVID-19 patients (n=16). (**D**) UMAPs on concatenated files (see Methods) of total neutrophils in healthy controls, moderate and severe COVID-19 patients. (**E**) UMAPs of CD16^bright^ and CD16^dim^ cells within pooled neutrophils from all study groups (30,000 down-sampled cells/individual). (**F**) Median fluorescence intensity (MFI) of selected surface markers in the UMAP projection described in (**D**). (**G**) Heatmap showing the mean normalized marker expression (*z-*score) in CD16^bright^ and CD16^dim^ neutrophils in healthy controls (n=17), moderate (n=10) and severe COVID-19 patients (n=16). (**H**) PCA based on marker expression (MFI) on CD16^bright^ and CD16^dim^ neutrophils. (**B)** and (**C**) Kruskall-Wallis test and two-stage Benjamini, Krieger and Yekutieli test. Bars represent median. * p < 0.05; ** p < 0.01; **** p < 0.0001.

A significant increase in absolute numbers was detected for both CD16^bright^ and CD16^dim^ neutrophil subsets in COVID-19 patients, which positively correlated with disease severity (Figure 2B). In addition, a significant increase in the relative fraction of CD16^dim^ neutrophils was detected, indicating an overall expansion of immature neutrophils in COVID-19 patients (Figure 2C).

In order to analyze the phenotypic traits of the neutrophil subsets identified above in an unsupervised way, 30,000 events in the neutrophil gate from each study participant were concatenated and then down-sampled to 0.5 million cells. By integrating the expression of 20 phenotypic markers in the UMAP analysis, we observed only modest differences comparing neutrophils from healthy controls and COVID-19 patients, within the UMAP space (Figure 2D). In addition, the CD16^bright^ and CD16^dim^ neutrophil subsets from controls and COVID-19 patients clustered separately from each other (Figure 2E), indicating substantial phenotypic differences, which did not depend on the disease state.

Next, the expression of markers related to neutrophil development, migration and activation was analyzed. The mature CD16^bright^ neutrophil subset was characterized by higher expression of CD177, CD11b, CXCR1, CXCR2 and CD62L compared to the CD16^dim^ population. On the other hand, CD16^dim^ neutrophils were characterized by higher levels of CD66b, LOX-1 and CD24, which are associated to early stages of neutrophil development (31), confirming the immature phenotype of this subset (Figures 2F-G, Supplemental Figures 2A-B).

We also observed significant upregulation of CD66b, CD177, CD11b, CXCR4, CD147 (the receptor that binds the spike protein of SARS-CoV-2) and CD63 in mature neutrophils from COVID-19 patients compared to healthy controls, as well as significant downregulation of CXCR2 (Figure 2G, Supplemental Figure 2A). No differences in the expression of markers related to either co-stimulatory or inhibitory functions, such as HLA-DR, CD86 and PD-L1 between healthy controls and COVID-19 patients within either neutrophil subset were detected (Figure 2G, Supplemental Figures 2A-B).

While the presented analysis showed several phenotypic differences between healthy controls and COVID-19 patients, we were unable to detect prominent alterations between the moderate and severe patients for any of the neutrophil subsets analyzed (Figure 2G, Supplemental Figure 2A-B). Such phenotypic similarity between the global neutrophil compartment in moderate and severe COVID-19 patients was confirmed by principal component analysis (PCA) (Figure 2H).

Collectively, our analysis highlights the accumulation of immature CD16^dim^ neutrophils and the concurrent activation of the mature neutrophil fraction in peripheral blood of COVID-19 patients.

### The abundance of activated neutrophil immunotypes correlates with markers of anti-viral immune response in moderate patients

To investigate further whether specific neutrophil phenotypes were associated with disease progression or severity, unsupervised phenograph clustering was performed on neutrophils derived from all study participants (Figure 3A). In total, 27 clusters were identified with a varying prevalence in the three study groups (Figure 3B). Hierarchical clustering based on marker expression was sufficient to group clusters more abundant in moderate patients (i.e. clusters 16, 22 and 27) (Figure 3C). Specifically, these clusters belonged to the CD16^bright^ subset and displayed higher expression levels of CD11b, CD177 and CD66b. On the other hand, clusters that were enriched in severe COVID-19 encompassed the immature CD16^dim^ population and displayed a more heterogeneous phenotype (Figure 3C). In particular, clusters 18, 19 and 23, which were more enriched in severe patients, showed lower levels of several activation markers including CD66b, CD11a and CD11b (Supplemental Figure 3A). Correlation of cluster frequencies in each patient with the concentration of 253 serum/plasma factors (Supplemental Table 4) followed by hierarchical clustering highlighted specific correlation patterns associated with trends in cluster enrichment within specific disease groups (Figure 3D). In particular, clusters 16, 22 and 27, which were enriched in moderate COVID-19, displayed a positive correlation with several molecules of type 1 response (CXCL9, CXCL10 and IL-15), anti-viral response (interferon regulatory factor 9 (IRF9), IL-12p70 and interferon-*γ* (IFN*γ*)) and adaptive immunity (B-cell activating factor (BAFF) and IL-2) (Figure 3D). In addition, key molecules involved in eosinophil/basophil homeostasis and recruitment (IL-5, CCL3 and stem cell factor (SCF)) were also positively correlated with these clusters. Within the clusters associated with severe patients, several members of the coagulation cascade (Factor VII, VIII and IX) and molecules associated with neutrophil maturation (i.e. granulocyte-colony stimulating factor (G-CSF) and myeloperoxidase (MPO)) were positively correlated (Figure 3D). Shared patterns within cluster groups (e.g. 22, 27 and 5, 23) were also observed when separately analyzing correlations in moderate and severe COVID-19 (Supplemental Figure 3B and Supplemental Table 5), suggesting that the degree of disease severity could influence relationships between specific neutrophil immunotypes and soluble factors.

**Figure 3.**
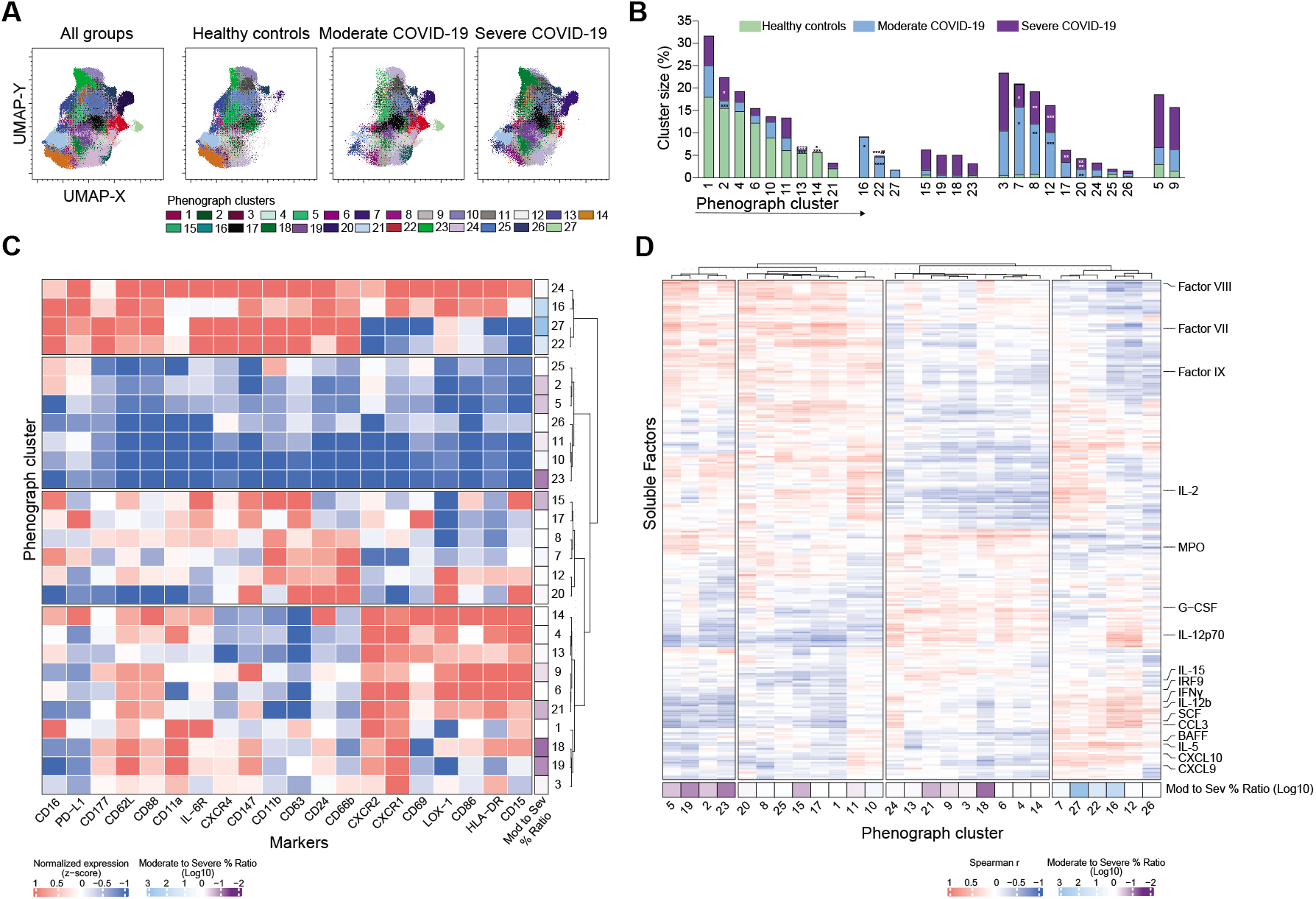
Phenograph analysis identifies activated neutrophil subsets correlating with markers of anti-viral immune response in moderate COVID-19 patients. (**A**) Distribution of the 27 identified phenograph clusters overlaid on the UMAP projection. (**B**) Frequency of the phenograph clusters within healthy controls (n=17), moderate (n=10) and severe COVID-19 patients (n=16). **\***, statistically significant differences in comparisons with healthy controls. **#**, statistically significant differences comparing moderate and severe patients. (**C**) Heatmap showing normalized expression (MFI *z*-score) of 20 markers analyzed by flow cytometry in the 27 neutrophil clusters identified in (**A**). Hierarchical clustering was performed and rows were split by *k*-means=4. Right annotation shows the log_10_-ratio of cluster enrichment in moderate (blue) versus severe (purple) COVID-19 patients. (**D**) Heatmap showing Spearman correlation between phenograph clusters and soluble factors detected in all COVID-19 patients (n=26). Hierarchical clustering was performed, and columns were split by *k*-means=4. Bottom annotation shows the log_10_-ratio of cluster enrichment in moderate (blue) versus severe (purple) COVID-19 patients. In (**B**) Kruskall-Wallis test and two-stage Benjamini, Krieger and Yekutieli test. * p < 0.05; ** p < 0.01; **** p < 0.0001.

In summary, the phenograph-guided in-depth dissection of the neutrophil subset composition allowed us to identify selected immunotypes associated with high circulating levels of prototypical molecules of type-1 related anti-viral immune response.

### Eosinophil activation profile in COVID-19 correlates with markers of anti-viral immune response

Through UMAP analysis integrating the expression of 21 surface markers on eosinophils, we observed that eosinophils from healthy donors clustered separately from the eosinophils of patients with COVID-19 (Figure 4A), indicating significant phenotypic alterations in the eosinophil populations upon SARS-CoV-2 infection. Eosinophils from COVID-19 had decreased expression of the markers of the eosinophil lineage (CD15, CD66b and CD193) (32) and higher expression of classical activation markers, such as CD62L, CD69, as well as CD147 (Figure 4B, Supplemental Figure 4A). Phenotypic analysis showed differences between healthy controls and COVID-19 patients, but also between patients with moderate and severe COVID-19 for some of the parameters analyzed. Patients with moderate COVID-19 expressed significantly higher levels of CD66b, CD11b, CD11a and CD24 compared to patients with severe COVID-19 (Figure 4C, Supplemental Figure 4B). Notably within the severe group, the deceased patients had the highest CD69 expression (Figure 4C). PCA analysis recapitulated these differences, highlighting CD193, CD66b, CD11a, CD24, CXCR4 and CD62L as the mostly affected parameters in eosinophils from COVID-19 patients (Figure 4D).

**Figure 4:**
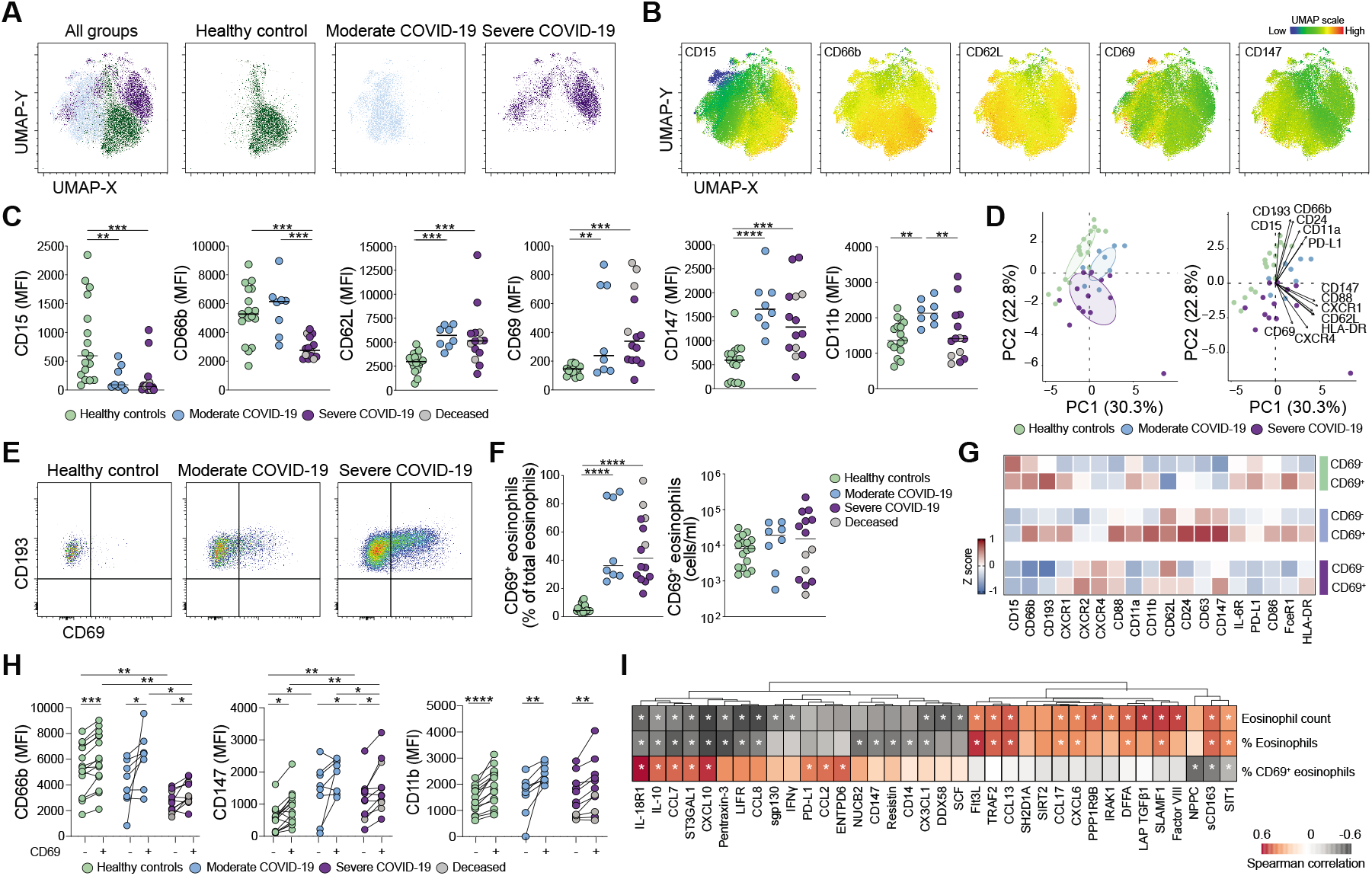
Activation of eosinophils in patients with COVID-19. (**A**) Overall and individual UMAPs on concatenated files (see Methods) of eosinophils in healthy controls, moderate and severe COVID-19 patients. (**B**) UMAP displaying the MFI of selected markers. (**C**) MFI of selected eosinophil markers in healthy controls (n=17), patients with moderate (n=8) and severe (n=14) COVID-19. Median values for each group are indicated. (**D**) PCA based on MFI expression of eosinophil markers. **(E)** CD69^+^ eosinophils from a representative healthy control and COVID-19 patients. (**F**) Frequencies and absolute counts of CD69^+^ eosinophils in healthy controls (n=17), moderate (n=8) and severe (n=14) COVID-19 patients. (**G**) Marker expression (*z*-score) on CD69^-^ and CD69^+^ subsets. (**H**) MFI of selected markers in eosinophil subsets in healthy controls (n=15), in moderate (n=8) and severe (n=11) COVID-19 patients. (**I**) Heatmaps demonstrating correlations (Spearman non-parametric, *r* < −0.4 or *r* > 0.4, *p* < 0.05) between absolute eosinophil counts, frequencies and percent of CD69^+^ eosinophils and soluble factors in COVID-19 patients. Statistical analysis by Kruskall-Wallis test and two-stage Benjamini, Krieger and Yekutieli test (**C, F** and **H)** and Wilcoxon matched-pairs signed rank test (**H**). FDR adjusted p-values are indicated. * p < 0.05; ** p < 0.01; *** p < 0.001; **** p < 0.0001.

We observed that patients with COVID-19 also had a significantly increased frequency of CD69^+^ eosinophils (Figures 4E-F). Indeed, despite the strong eosinopenia (Figure 1D), the absolute counts of CD69^+^ eosinophils were not significantly altered in COVID-19 patients (Figure 4F). CD69^+^ eosinophils displayed significantly higher expression of CD66b, CD147, CD11b and CD193, compared to the corresponding CD69^-^ counterpart, confirming their activation state (Figures 4G-H, Supplemental Figure 4D). Despite their CD69 expression, activated eosinophils from severe COVID-19 patients displayed lower CD11a, CD66b, and CD147 expression compared to CD69^+^ eosinophils from the group with moderate disease (Figures 4G-H, Supplemental Figure 4D), suggesting a partial functional impairment of eosinophils in the more severe COVID-19 stages.

The absolute counts of eosinophils in blood correlated inversely with the peripheral levels of SCF and molecules involved in cell response to viral infection, such as DDX58 (DExD/H-Box Helicase 58, also known as RIG-I), IFN*γ* and CXCL10 (Figure 4I). Conversely, the eosinophil levels in blood correlated positively with the levels of Interleukin-1 receptor-associated kinase 1 (IRAK1), latency associated peptide-transforming growth factor β1 (LAP-TGFβ1), chemoattractants CCL13, CCL17 and CXCL6 and the coagulation factor VIII (Figure 4I). The frequency of CD69^+^ eosinophils correlated positively with the levels of molecules related to IFN*γ*, CCL2, CCL7, CCL8, Pentraxin-3, leukemia inhibitory factor receptor (LIFR), as well as PD-L1. sCD163 correlated negatively with the frequency of CD69^+^ eosinophils but positively with the absolute counts and frequencies of eosinophils. Representative plots of the most significantly correlated parameters are shown in Supplemental Figure 4E, while the most significant correlations for the moderate and severe groups separately are shown in Supplemental Figure 4F. In conclusion, our results indicate that the eosinophils in the blood of patients with COVID-19 are activated in response to SARS-CoV-2 infection and express high levels of key receptors for lung tissue infiltration, particularly in patients with moderate COVID-19.

### Basophils in COVID-19 patients display an activated phenotype

UMAP analyses revealed distinctive clusters in the healthy controls and COVID-19 patients, suggesting alterations in basophil phenotype during the disease (Figure 5A). Specifically, clusters associated with COVID-19 had higher expression of markers important for the activation and recruitment of basophils, including CD11b, CD63, and CXCR4 (Figure 5B; Supplemental Figure 5A). Specific upregulation of CD62L, CD147 and CD177 were observed on basophils from moderate COVID-19 patients (Figure 5C, Supplemental Figure 5B). On the other hand, PD-L1 expression was reduced in all patients compared to healthy controls. The differences in marker expression on basophils between healthy controls and COVID-19 patients were further confirmed by PCA, with CXCR4, CD62L, CD193 and PD-L1 among the most contributing parameters driving the separation (Figure 5D). We further examined the correlation of basophils with soluble factors (Figure 5E, Supplemental Figure 5D-E). The basophil frequency was negatively correlated with soluble DDX58, and with the transcription factors amphiregulin (AREG), FOSB and Zinc finger and BTB domain-containing protein 16 (ZBTB16). Similar to eosinophils, basophil frequency was also negatively correlated with CCL7. In contrast, CCL22, CCL28 and Fms-related tyrosine kinase 3 ligand (Flt3L) were positively correlated to basophil frequency, particularly in COVID-19 patients with moderate disease (Figure 5E, Supplemental Figure 5E). Other soluble factors that were also positively associated with basophil frequency were CD83 and tumor necrosis factor-related apoptosis-inducing ligand (TRAIL). In summary, the data show that basophils are depleted in COVID-19 patients, a pattern associated with disease severity, and that circulating basophils from patients display an activated phenotype.

**Figure 5.**
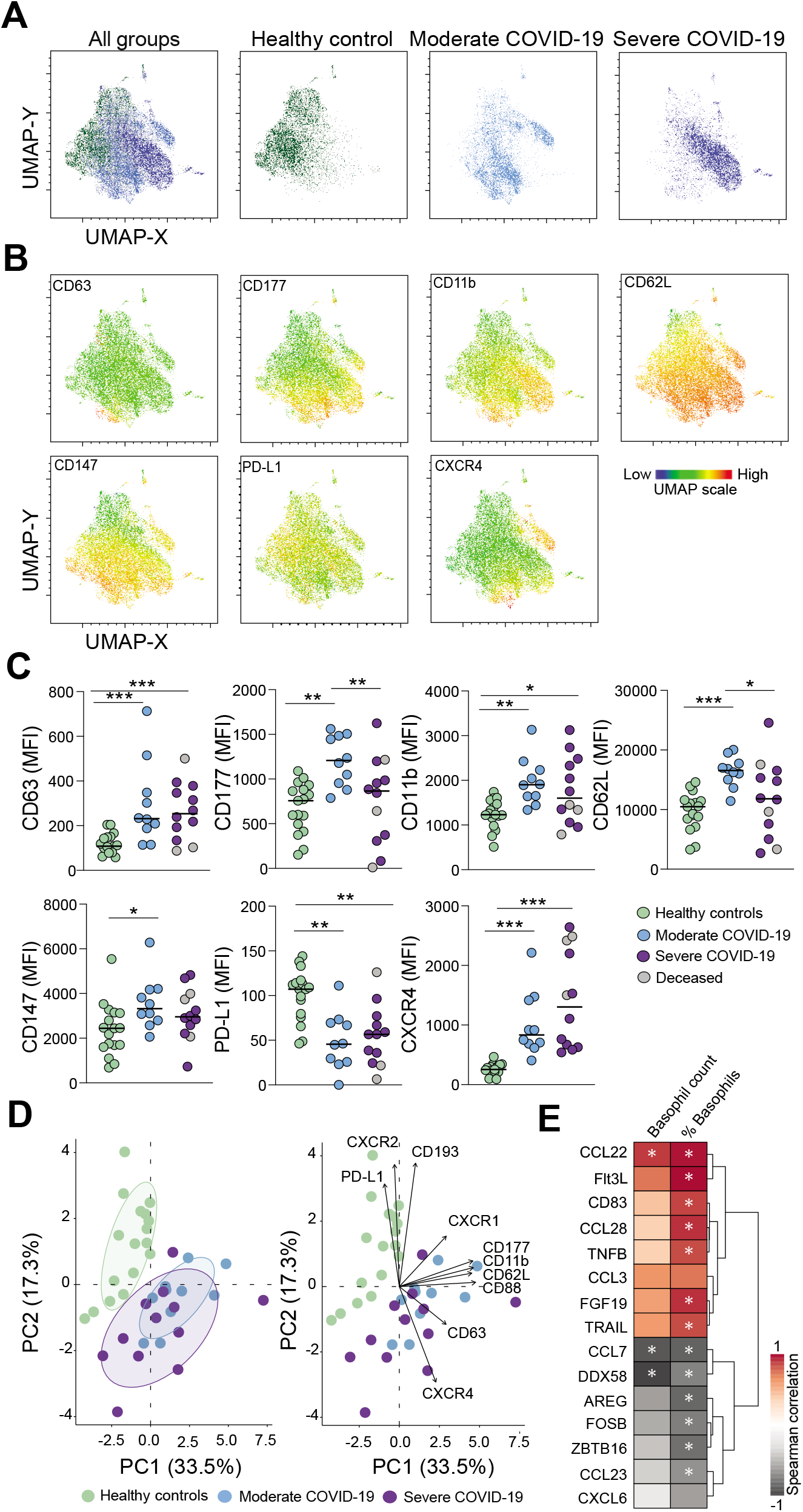
Basophils in COVID-19 patients display an activated phenotype. (**A**) UMAPs on concatenated files (see Methods) showing basophil overview together with representative plots heathy controls, moderate and severe COVID-19 patients. (**B**) UMAP showing the level of expression of selected markers on basophils. (**C**) MFI for selected markers on basophils in heathy controls (n=17), moderate (n=10) and severe (n=12) COVID-19 patients. Median values for each group are indicated. (**D**) PCA and biplot based on the MFI expression of representative basophil markers in healthy controls, moderate and severe COVID-19 patients. **(E)** Heatmap demonstrating correlation (Spearman non-parametric, *r* < −0.4 or *r* > 0.4, *p* < 0.05) between absolute basophil counts/basophil frequencies of total leukocytes and basophil-associated soluble factors, in COVID-19 patients. Significant differences between healthy controls and patient groups in (**C**) were evaluated with Kruskall-Wallis test and two-stage Benjamini, Krieger and Yekutieli test. FDR adjusted p-values are indicated. * p < 0.05; ** p < 0.01; *** p < 0.001.

### Eosinophil activation and neutrophil maturation contribute to predictive models of Sequential Organ Failure Assessment (SOFA) score and respiratory function

Our well-defined patient cohort allowed us to compare in detail the phenotypic alterations in granulocyte subsets in relation to severity of COVID-19 (Supplemental Table 1). In order to elucidate the underlying relationships between immune traits and relevant clinical parameters registered during hospitalization, we performed multivariate linear regression. The models were based on a comprehensive data set including surface marker expression levels, absolute numbers, frequencies of granulocyte subsets, available clinical information and laboratory measurements. The final set of variables used in the predictive models were selected based on their correlation with the tested clinical outcomes and their relevance in the specific predictive models (Figure 6A).

**Figure 6.**
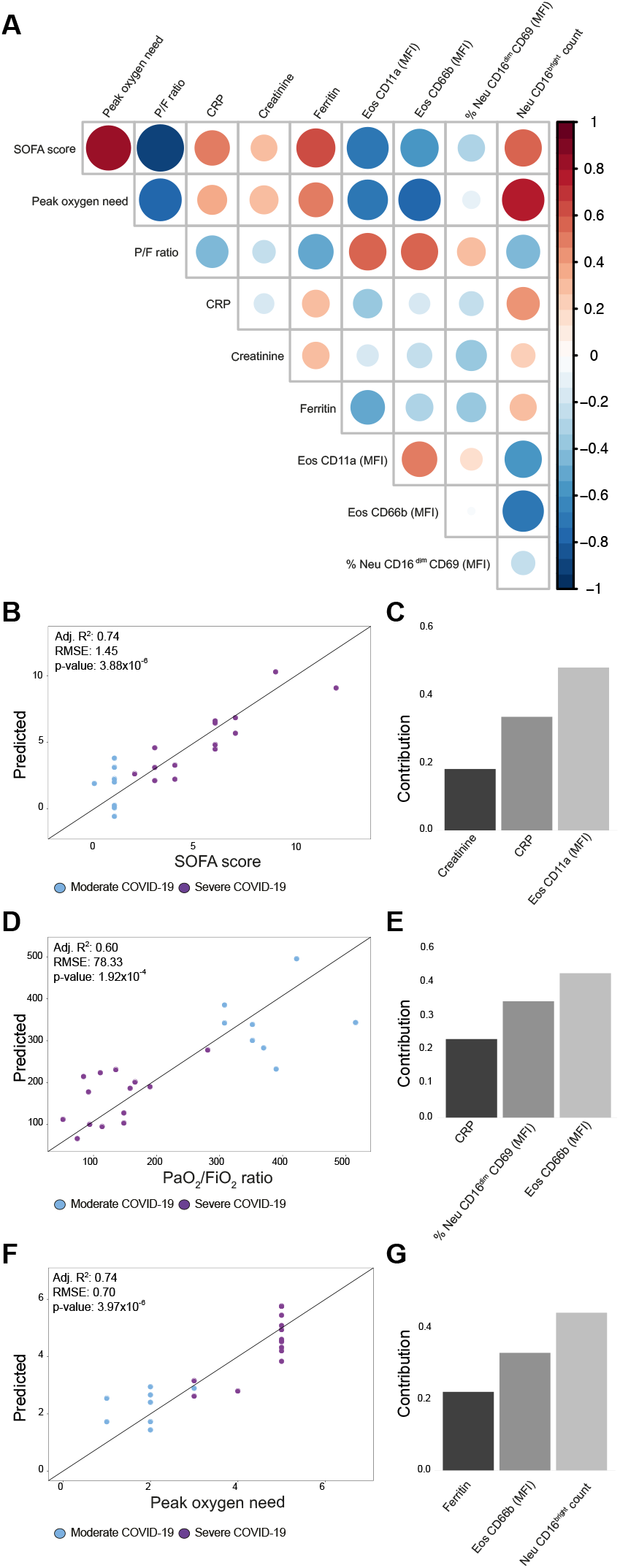
Eosinophil activation and neutrophil maturation are relevant for prediction of SOFA score and respiratory function. (**A**) Diagonal correlation matrix including predicted clinical outcomes (bold) and the explanatory variables included in the final models. (**B**-**G**) Multivariate linear regressions for prediction of SOFA score (**B**-**C**), PaO_2_/FiO_2_ (P/F) ratio (**D**-**E**) and peak oxygen need (**F**-**G**) were modeled based on immune traits and laboratory markers. (**B, D, F**) Scatter plots of actual values versus predicted values of the linear models. (**C, E, G**) Individual contribution of the variables included in the final models. Adj R^2^, Adjusted R^2^; RMSE, Root-mean-square error; Eos, Eosinophils; Neu, Neutrophils.

The analysis revealed that the levels of CD11a expression on eosinophils contributed to the linear model for prediction of SOFA score for patients included in our study (Adj. R^2^=0.74, p=3.8×10^−6^, Figure 6B). Notably, the relevance of CD11a expression on eosinophils was higher in predicting the SOFA score (explaining around 50% of the model) than the levels of two laboratory parameters CRP and creatinine (Figure 6C).

We also developed models for prediction of respiratory function including PaO_2_/FiO_2_ (P/F) ratio at baseline and maximum oxygen need during hospitalization. The linear model for P/F ratio prediction (Adj. R^2^=0.60, p=1.9×10^−4^, Figure 6D) included contributions of CD69 expression on CD16^dim^ neutrophils, of CD66b on eosinophils and of CRP. The linear model for maximum level of oxygen administration encompassed absolute numbers of mature neutrophils, CD66b expression on eosinophils and ferritin levels (Adj. R^2^=0.74, p=3.9×10^−6^, Figure 6F). Notably, both models revealed a correlation between CD66b expression on eosinophils and degree of respiratory failure during COVID-19. Moreover, the combined effect of the immune parameters in both models was dominant, contributing to almost 80% of each model (Figures 6C, 6E and 6G). Correction for relevant parameters (e.g. age, BMI, sex, co-morbidities) did not significantly alter any of the linear models or the contribution of the immune cell populations to them, further strengthening the link between granulocyte activation and clinical outcome (Supplemental Table 6). In summary, by applying a supervised machine-learning method we identified eosinophil activation and mature neutrophil counts to be strongly correlated to SOFA score and maximum oxygen need. These immunological signatures, together with other known laboratory markers, but not alone (Supplemental Figure 6), were sufficient to create predictive models.

### The phenotype of granulocytes is partly restored in patients who have recovered from COVID-19

To determine whether the phenotypical alterations observed within the granulocyte compartment during acute COVID-19 were recovered after viral clearance, whole blood samples were collected from the same moderate (n=8) or severe (n=7) patients approximately four months (median=136 days, range=89-153 days) after hospital discharge (Figure 7A). Convalescent samples displayed normalized cell counts for neutrophils, eosinophils and basophils, as well as neutrophil-to-lymphocyte ratio, compared to the samples collected during acute viral infection (Figure 7B, Supplemental Figure 7A), while the relative abundance of the granulocyte subsets over total leukocytes was only partially restored (Figure 7C), suggesting that circulating non-granulocytic immune cells might still be quantitatively affected. Nevertheless, both absolute numbers and frequencies of granulocyte subsets in convalescent patients were very similar to those observed in healthy controls (Figures 7B-C). Additionally, PCA highlighted that convalescent patients were markedly more similar to healthy controls than to acute patients (Figure 7D). In particular, CD69 and CD193 expression on eosinophils, as well as CD147 expression on neutrophils and eosinophils, and basophil counts were among the variables contributing to the observed clustering (Figures 7E-F, Supplemental Figure 7B).

**Figure 7.**
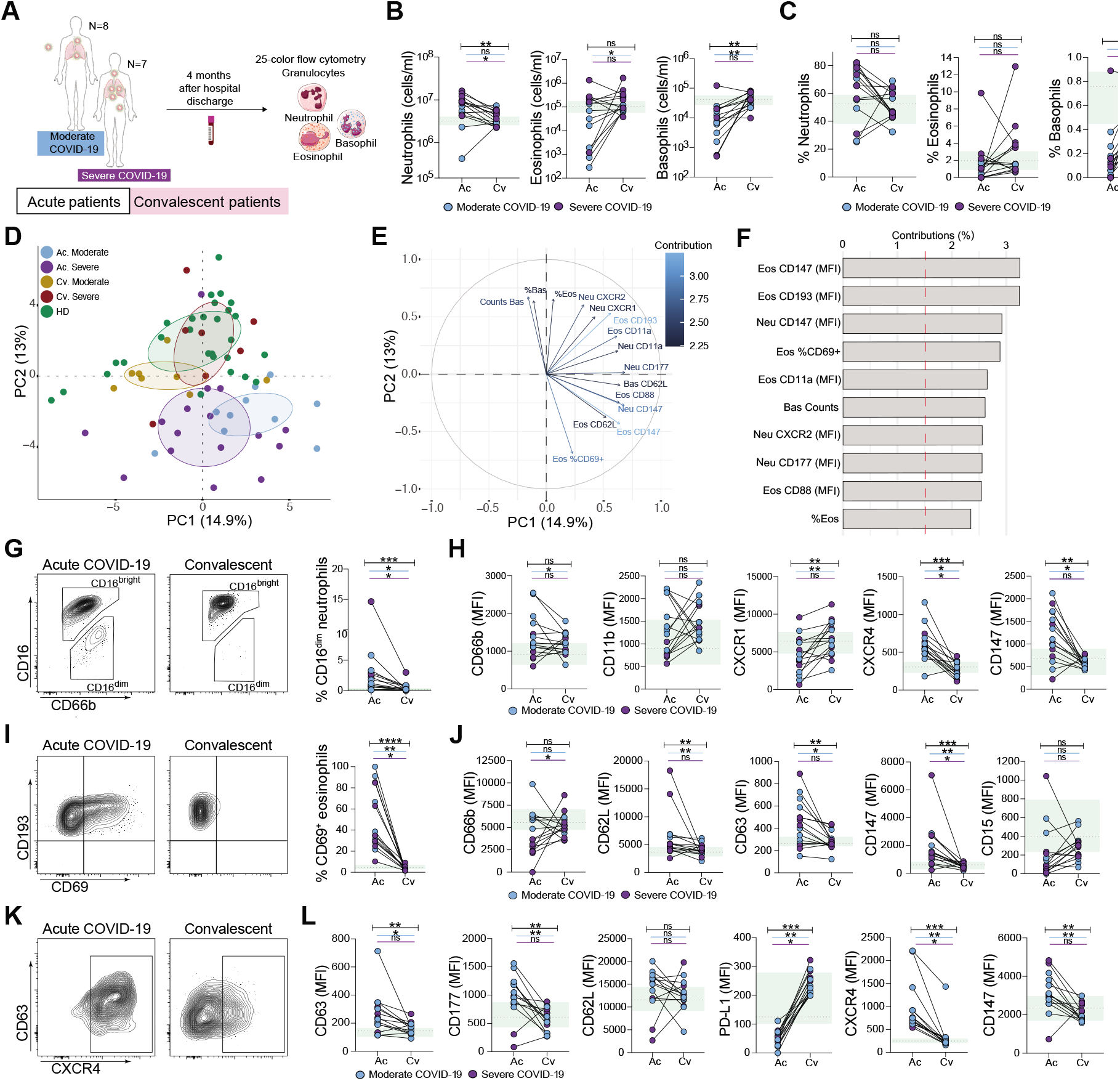
Granulocyte phenotype in convalescent patients is largely restored. (**A**) Schematic illustration of paired acute (Ac) and convalescent (Cv) patient samples included in the study. Absolute cell counts (**B**) and granulocyte frequencies over total leukocytes (**C**) in paired Ac and Cv patients (n=15). (**D**) PCA based on cell counts, frequencies and normalized MFI (*z*-score) of all measured markers. (**E**) Variable correlation plot. (**F**) Relative contribution of variables to PC1 and PC2. (**G**) CD16^dim^ neutrophil frequencies in paired Ac and Cv patient samples (n=15). (**H**) Expression of selected neutrophil markers in paired Ac and Cv patients (n=15). (**I**) CD69 eosinophil expression in paired Ac and Cv patients (n=15). (**J**) Expression of selected eosinophil markers in paired Ac and Cv patient samples (n=15). (**K**) CXCR4 basophil expression in Ac and Cv patients. (**L**) Expression of selected basophil markers in paired Ac and Cv patients (n=15). Wilcoxon matched-pairs rank test; ns, not significant; * p < 0.05; ** p < 0.01; *** p < 0.001; **** p < 0.0001. Healthy controls (n=28, median±IQR) ranges are shown in green. Bars indicate statistical significance considering all sampled patients (black), only moderate (blue) or only severe (purple) patients. Bas, Basophils; Eos, Eosinophils; Neu, Neutrophils.

Immature CD16^dim^ neutrophils, elevated in acute COVID-19, were almost absent in convalescence (Figure 7G). Neutrophils from convalescent patients showed normalized levels of CXCR1, CXCR4 and CD147 compared to acute infection (Figure 7H, Supplemental Figure 7C). Eosinophil activation was also reduced in convalescent patients, as shown by the normalized expression of CD69, CD62L, CD63, and CD147, as well as by the increased PD-L1 expression (Figures. 7I-J, Supplemental Figure 7D). However, the expression of CD66b, CD15 and CD193 on eosinophils was not completely recovered in convalescent samples when compared to healthy controls (Figure 7J, Supplemental Figure 7D), suggesting that a partial phenotypic impairment might still be present in the eosinophil population. Similarly, basophils in the convalescent samples showed an overall recovery of several phenotypic traits altered in acute COVID-19. In particular, the expression levels of CD11b, CD63, CXCR4, and PD-L1 (Figures 7K-L, Supplemental Figure 7E) were in line with the levels found in healthy controls.

## Discussion

The reported clinical relevance of the inflammatory state in the more severe forms of COVID-19 has raised an unprecedented awareness on the potential detrimental role of polymorphonuclear cells in acute viral infections (19). However, a comprehensive phenotypic description performed at the protein level on circulating granulocytes in SARS-CoV-2-infected patients is still lacking. In this report, we show that immature CD16^dim^ neutrophils accumulate in peripheral blood of COVID-19 patients and confirm that general neutrophilia can be considered a hallmark of severe COVID-19. However, in contrast to the tenet linking neutrophil activation with more severe clinical conditions, our approach based on high-dimensional flow cytometry was able to identify activated neutrophil immunotypes detected preferentially in moderate patients showing a better clinical outcome. Furthermore, we extended our analysis to other granulocyte subsets, reporting the substantial ablation of both eosinophils and basophils in peripheral blood of COVID-19 patients. Such depletion could possibly result from two, non-mutually exclusive phenomena: first, both cell types might be recruited to inflamed tissues, and in particular to the lung. This hypothesis is supported by the relevant expression changes of several adhesion/migration molecules (e.g. CD62L, CD11a/b and CXCR4) on both eosinophils and basophils. Moreover, concentrations of key soluble factors involved in the recruitment of both granulocyte subsets (e.g. CCL13, CCL17, CCL22 and CCL28) (33) correlated with their levels in circulation. A second explanation for the detected basopenia/eosinopenia in favor of an expanded neutrophil compartment might imply the re-programming of the granulocyte precursor in the bone marrow which typically occurs during emergency hematopoiesis (34).

The release of immature neutrophils in circulation has been observed in previous studies investigating a variety of viral infections, including HIV, dengue and SARS-CoV-2 (18, 19, 35-38) and is strictly associated with emergency granulopoiesis. Recent studies included CD63 mRNA levels in an immature pro/pre-neutrophil signature (31), which was found significantly enriched in COVID-19 patients (19). In this regard, we found a robust CD63 upregulation on CD16^dim^ neutrophils and a further increased expression in COVID-19 patients. Both moderate and severe COVID-19 patients were characterized by the progressive expansion of CD16^dim^ neutrophils, which also expressed lower levels of CD177, CD11b and CD62L and higher levels of CD66b and LOX-1, a phenotype compatible with neutrophil immaturity (31).

Our in-depth phenograph-guided analysis of the neutrophil compartment revealed the enrichment of activated neutrophil immunotypes in moderate COVID-19 patients, associated with higher circulating levels of molecules involved in the IFN-mediated anti-viral response (i.e. IRF9, IL-12b, IFN*γ*). Neutrophils are known to be highly responsive to type I IFN in different contexts (39, 40) and recent studies described the existence of an IFN-responsive, developmentally distinct, neutrophil subset (41). Intriguingly, we observed a positive correlation between type 1 inflammatory mediators (e.g. IFN*γ* and CXCL10) and eosinophil activation, suggesting that, particularly in moderately affected patients, part of the granulocyte compartment could be actively participating in the efficient viral clearance, similarly to what occurs upon influenza infection (42, 43). Interestingly, CD62L expression on eosinophils can be triggered by IFN*γ*, and CD62L-expressing eosinophils have been suggested to contribute to dysregulated inflammation and ARDS in acute COVID-19 (24, 44).

Eosinophils have been reported to express receptors that allow the recognition and orchestration of anti-viral responses to respiratory viruses (43, 45, 46). Eosinophil-associated lung pathology has been reported in other viral infections and in SARS-CoV-1 vaccination studies (summarized in (47)). On the other hand, the role of human basophils in viral infections is poorly understood and mainly focused on basophil response to HIV (48, 49). CXCR4 is one of the most highly expressed basophil receptors in COVID-19 patients in our cohort and might be implicated in basophil trans-endothelial migration (49). CD63 expression on basophils can be induced by cross-linking of CD62L and CD11b, amongst other stimuli (50). Therefore, the upregulation of CD62L, CD63, CD11b, and CXCR4 on basophils observed during the acute phase of COVID-19 and their normalization after viral clearance might imply a role of this phenotype in COVID-19 pathophysiology.

In contrast to the neutrophil phenotypes observed in moderate COVID-19, the neutrophil clusters enriched in patients with severe disease correlated with several coagulation factors and with molecules involved in neutrophil maturation (G-CSF and MPO), but not with any of the screened anti-viral response-related molecules. This suggests an impaired neutrophil response during the more severe phases of the disease. Notably, we did not detect evident signs of immune-suppressive activity (e.g. PD-L1 expression) in our phenotypic analysis, and direct functional evidence will be crucial to assess whether increased suppressive functions are triggered in neutrophils in severe COVID-19 patients. In addition, despite their CD69 expression, activated eosinophils in severe COVID-19 patients displayed lower levels of CD11a, CD63, and CD66b compared to the CD69^+^ eosinophils from patients with moderate COVID-19. This phenotype is indicative of activated eosinophils suggesting that they might have recently degranulated (32, 51). Overall, our analyses showed that the robust activation observed in the granulocyte compartment in moderate COVID-19 declines in the more severe stages of the disease.

Combination of eosinopenia with elevated CRP could effectively triage suspected patients with COVID-19 from other patients with fever (52). A relevant finding emerging from our study is that selected granulocytic phenotypical traits could, in concert with other known laboratory markers such as CRP or creatinine, predict to a significant extent, key clinical outcomes, including respiratory functionality and SOFA score. Although this finding needs to be taken with caution considering the limited size of our patient cohort, we provide the proof-of-principle that the combination of immunological, and specifically granulocyte-related, measurements with standard clinical data could be used in generating algorithms for patient classification and, thus, tailored therapeutic regimens. Notably, the immunological parameters were predominant in driving the prediction models in comparison to standard laboratory measurements. On the same lines, we connected clinical manifestations, such multi-organ failure and pulmonary function, with specific phenotypes, primarily the eosinophil activation markers CD11a, CD66b, and CD69. Future studies will be important to assess and validate whether there is a mechanistic link between granulocyte activation and specific clinical features of COVID-19 patients.

Finally, by taking advantage of paired longitudinal sampling, we report the almost complete phenotypic recovery upon viral clearance of the phenotypic alterations observed in granulocytes from acute SARS-CoV-2 infection. Our findings are in agreement with previous studies showing the replenishment of the eosinophil and basophil pool and the normalization of neutrophil numbers in circulation (24, 53). The current study, however, is the first to characterize in depth the eosinophil and basophil profile in patients with COVID-19 and their phenotypic alterations linked to disease. This is emphasized by the finding that six out of the ten most significant immunological parameters that drive the separate clustering of patients with acute COVID-19 from matched convalescent samples or healthy controls are eosinophil traits. However, the expression of a set of markers (CD15, CD66b, and CD193) was not completely recovered in convalescent samples when compared to non-infected healthy controls, suggesting a partial remaining phenotypic impairment in the granulocyte population in convalescent samples, at least four months after hospital discharge.

The inference of mechanisms driving COVID-19 immunopathology from the analysis of circulating granulocytes might have limitations when considering the pulmonary damage displayed in COVID-19 patients. However, several studies have shown that the combined use of peripheral immune signatures, soluble factors and patient metadata retain the capacity to predict clinical outcome to a notable extent (6, 53). Moreover, accumulating evidence indicates that other tissues (e.g. kidney, gut, brain) are affected by COVID-19-related immune alterations, thus underscoring the systemic nature of this disease and emphasizing the relevance of studying alterations in peripheral immune cells (54). Our findings highlight the significant alterations of granulocyte subpopulations in frequency and function in the blood of patients with COVID-19. Moreover, our data indicate the potential contribution of granulocytes to SARS-CoV-2 immunopathology and point towards the combined use of granulocyte-related immunological parameters and basic clinical laboratory tests as better prognostic biomarkers of disease severity and disease course.

## Materials and Methods

### Study cohort

SARS-CoV-2 infected patients with COVID-19 admitted at the intensive care or high dependency unit (n=16, severe COVID-19) or the infectious disease clinic (n=10, moderate COVID-19) at Karolinska University Hospital, Stockholm, Sweden, were recruited to this study. Serum and EDTA blood samples were collected and analyzed. Inclusion and exclusion criteria for patient enrollment and collected clinical information and laboratory values of the patients are provided in Supplemental Tables 1-3. Convalescent serum and EDTA samples were later collected from 15 patients (8 and 7, from the moderate and severe groups, respectively) approximately 4 months after hospital discharge (median=136 days, range=89-153 days). As controls, serum and blood samples from age- and sex-matched SARS-CoV-2 IgG seronegative healthy volunteers (Supplemental Table 1) were collected on the same days as the acute COVID-19 (n=17) and convalescent (n=11) patients. All samples were processed using the same standardized operating procedures. Informed consent was obtained from all study participants and the study was approved by the regional Ethics Committee in Stockholm, Sweden, and performed in accordance with the Declaration of Helsinki.

### Absolute counts of leukocytes in peripheral blood

Absolute counts of peripheral blood leukocytes were determined for all study participants using BD Trucount tubes (6-color TBNK Reagent and, in addition, CD123 BUV395 (BD Biosciences), CD15 PB, CD193 BV605 and HLA-DR BV785 (Biolegend) and CD14 PE-Cy5 (eBioscience) according to the manufacturer’s instructions. Samples were fixed in 1X BD FACS lysing solution (BD Biosciences) for 2h and acquired on a BD FACSymphony A5 instrument, equipped with UV (355nm), violet (405 nm), blue (488 nm), yellow/green (561 nm) and red (637 nm) lasers. For absolute cell count calculations, the number of events for populations of interest was divided by the number of bead events and multiplied by the BD Trucount bead count.

### Proximity extension assay

The proximity extension assay technology (PEA), based on Q-PCR quantification of pair-wise binding of oligonucleotide-labeled target antibodies (OLINK AB, Uppsala, Sweden), was used for the quantification of 276 selected soluble factors in serum from the study participants (Supplemental Table 4).

### Luminex assays

Soluble factors in the serum of the study participants were measured using 4 customized multiplex panels including 36, 12, 15 and 12 analytes covering a range of human inflammatory factors (R&D Systems, UK; Supplemental Table 4). Coagulation factors were measured in plasma using 3 different multiplex panels, including the 6-, 4-, and 3-plex human ProcartaPlex panels (ThermoFisher). Assays were performed according to the manufacturer’s guidelines and samples were acquired on a Luminex MAGPIX instrument using xPonent 4.0 software (Luminex).

### Cell preparation and staining for multicolor flow cytometry

For flow cytometric analysis of polymorphonuclear leukocytes, EDTA whole blood samples from all patients and controls were stained with fluorescently labeled antibodies (Supplemental Table 7). In brief, plasma fractions were removed by centrifugation and cells were washed with FACS buffer (PBS, 5% FCS, 0.05 mM EDTA), followed by 15-minute incubation with a cocktail containing Fc-block (Miltenyi) and fluorescent-labeled antibodies for extracellular staining. Dead cells were excluded using the fixable LIVE/DEAD Yellow Dead Cell Stain Kit

(Life Technologies). Red blood cells (RBC) were lysed with fixation/permeabilization working solution (BD Cytofix/Cytoperm™). For SARS-CoV-2 inactivation, samples were fixed for two additional hours in ultra-pure, methanol-free, 1% formaldehyde (Polyscience). Due to heavy RBC coagulation in COVID-19 patient samples, an additional RBC lysis for 10 min was performed. Samples were acquired on a BD FACSymphony equipped with five lasers (BD Biosciences).

### Flow cytometry data analysis

Standard flow cytometry data analysis was performed using FlowJo version 10. A compensation matrix for the 25-color flow cytometry panel was generated using AutoSpill (55), optimized and applied to all fcs files. All data was pre-processed using the time gate. Following exclusion of doublets and dead cells, neutrophils were defined as CD15^+^CD193^-^ CD66b^+^CD16^bright/dim^, eosinophils were defined as CD15^bright/dim^CD193^bright/dim^CD16^-^ FcER1^dim^CD123^dim/-^ and basophils were defined as CD15^-^ CD193^bright^FcER1^bright^CD123^bright^HLA-DR^dim/-^. In addition, exclusion of potential contamination by non-granulocytes was done using lineage exclusion markers (CD3, CD14, CD19, CD56, CD304). Samples containing less than 30 events in the final granulocyte subset gate were not considered for further phenotypical analysis. The Cytonorm FlowJo plugin v1.0 was applied on each granulocyte population to correct for potential batch effects and was based on an internal control that was sampled on all sampling occasions. Dimensionality reduction was performed with the UMAP FlowJo plugin v3.1. Neutrophil populations from each individual were down-sampled for comparability (FlowJo Downsample plugin v3.3), barcoded and concatenated. FlowJo Phenograph v3 was used for unsupervised clustering.

### Statistical analysis

GraphPad Prism version 9 (GraphPad Software) and R v4.0.1 (56) were used to conduct statistical analyses, where p-values < 0.05 were considered significant. Two-tailed and non-parametric Mann-Whitney *U* test was used for two-group comparisons. Non-parametric Kruskal-Wallis test, in combination with two-stage step-up method of Benjamini, Krieger and Yekutieli for controlling False Discovery rate (FDR), was used for multiple-group comparisons. Paired samples were analyzed with Wilcoxon matched-pairs signed rank test. For correlations, the two-tailed non-parametric Spearman test was applied. Where indicated, *Ζ*-score of median fluorescence intensity (MFI) was calculated as follows: *z* = (*x*-*μ*)/*σ*, being *x* = raw score, *µ* = mean of sample distribution and *σ* = standard deviation. Correlation, hierarchical clustering and multivariate analysis of flow cytometry data with clinical parameters and proteomic data was performed and visualized in GraphPad Software and R v4.0.1 and v1.3.959 (56), using the packages factoextra (v1.0.7) (57), FactoMineR (v2.3) (58), PerfomanceAnalytics (v2.0.4) (59), ggplot2 (v3.3.1) (60), gplots (v3.0.4) (61), pheatmap (v1.0.12) (62), vegan (v2.5-6) (63), corrplot (v0.84) (64), lattice (v0.20-41) (65) and latticeExtra (v0.6-29) (66), stats (v4.0.1) and complexheatmap (v2.5.6) (67).

### Multivariable linear regression

Predictive models for clinical outcomes were built based on a data set of 167 immune traits that included surface marker expression levels, absolute numbers, frequencies of granulocyte subsets, available clinical information and laboratory measured parameters. The creation of the models was performed in R v. 3.6.0 (56). Variable redundancy was avoided by identification of variables with pair-wise spearman correlation coefficient higher than 0.99 (function cor from package stats), and removal of the variable with the largest mean absolute correlation. The remaining variables were pair-wise correlated with the clinical outcome, and those with p-values < 0.05 were selected for further evaluation. All possible linear combinations of variables were evaluated with the function regsubsets from the package leaps (68) using an ‘exhaustive’ method and evaluating 3 subsets per model size. The models with highest adjusted R^2^, lowest bayesian information criterion and a Mallow’s Cp lower and closer to the number of coefficients, were selected for further evaluation. The variance inflation factor for each model was calculated to avoid redundancy in the independent variables with the function vir from package car (69). The selected final model had no redundant variables, and all must be significant (p-value < 0.05). The contribution of each parameter to each model was calculated with the functions cal.relimp from package relaimpo (70) with the lgm metric. Differences in the model after addition of factors for correction (sex, age, BMI, smoking, days from symptom to sampling and from admission to sampling and co-morbidities) were evaluated by computing an analysis of variance for the two linear models.

## Data Availability

Curated flow cytometry data will be made available for exploration via the KI/K COVID-19 Immune Atlas, http://covid19cellatlas.com/. All other data needed to evaluate the conclusions of the paper are presented in the paper or in the Supplemental Material. 

## Author contributions

Conceptualization: ML, MD, LH, AP, ANT, BJC. Methodology: ML, MD, LH, LMPM. Investigation: ML, MD, LH, LMPM, PC, JRM, J-BG, JE, KMo, MG, KMa. Data analysis: ML, MD, LH, HB, LMPM, EK, MC, AP. Patient recruitment and clinical information: EF, SGR, AS, LIE, OR, SA, KS, H-GL, NKB. Organization of sample handling and other resources: PC, JK, JMi, MFT, SB, MB, JMj, K-JM, JS, J-IH and the Karolinska KI/K COVID-19 Study Group. Writing (original draft): ML, MD, LH, HB, LMPM, AP, BJC, ANT. Writing (review and editing): All authors. Supervision of experiments: ML.

## Acknowledgements

We thank all patients and healthy donors for their generous participation.

See Supplemental Acknowledgments for Karolinska KI/K COVID-19 consortium details. The study was supported by grants from Nordstjernan AB and Knut and Alice Wallenberg Foundation to H-GL, the Swedish Governmental Agency for Innovation Systems (VINNOVA) under the frame of NordForsk (Project no. 90456, PerAID) and the Swedish Research Council under the frame of ERA PerMed (Project 2018-151, PerMIT) to ANT, the Swedish Research Council to BJC and AP, the Swedish Cancer Foundation to BJC, Clas Groschinsky Foundation, Centrum for Innovative Medicine to AP and ANT, the Swedish Children’s Cancer Foundation (TJ2018-0128 and PR2019-0100) to ML, Åke Olsson Foundation to ML and AP and KI Research Foundation to ML, LH and AP.

