## Supplemental material for "High-dimensional profiling reveals phenotypic heterogeneity and disease-specific alterations of granulocytes in COVID-19"

**Authorship note**:

ML, MD and LH share first authorship.

AP, ANT and BJC share last authorship.

**Affiliations**:

[1]Center for Infectious Medicine, Department of Medicine Huddinge, Karolinska Institutet, Karolinska University Hospital, Stockholm, Sweden

[2]Childhood Cancer Research Unit, Department of Women's and Children's Health, Karolinska Institutet, Stockholm, Sweden

[3]Theme of Children's Health, Karolinska University Hospital, Stockholm, Sweden

[4]Department of Infectious Diseases, Karolinska University Hospital, Stockholm, Sweden

[5]Division of Infectious Diseases, Department of Medicine Solna, Karolinska Institutet, Stockholm, Sweden

[6]Division of Infectious Diseases and Dermatology, Department of Medicine Huddinge, Karolinska Institutet, Stockholm, Sweden

[7]Department of Physiology and Pharmacology, Section for Anesthesiology and Intensive Care, Karolinska Institutet, Stockholm, Sweden

[8]Function Perioperative Medicine and Intensive Care, Karolinska University Hospital, Stockholm, Sweden

[9]Department of Clinical Interventions and Technology CLINTEC, Division for Anesthesiology and Intensive Care, Karolinska Institutet, Stockholm, Sweden.

### Supplemental Material

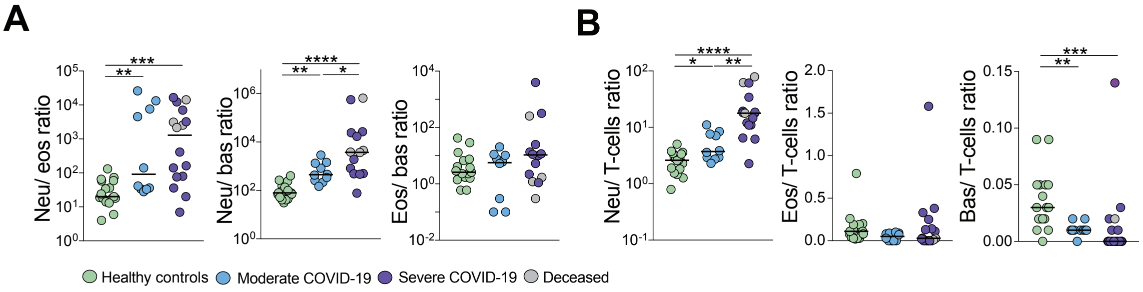

**Supplemental Figure 1**. **Granulocyte cell count ratios in COVID-19.** (**A**) Cell count ratios of granulocyte subsets (neu, neutrophils; eos, eosinophils; bas, basophils) and (**B**) of granulocyte subsets versus T-cells in healthy controls (n=17), moderate COVID-19 patients (n=10) and severe COVID-19 patients (n=16). Significant differences were evaluated with Kruskall-Wallis test and two-stage Benjamini, Krieger and Yekutieli test. FDR adjusted p-values are indicated**.** * $p < 0.05$; ** $p < 0.01$; *** $p < 0.001$; **** $p < 0.0001$.

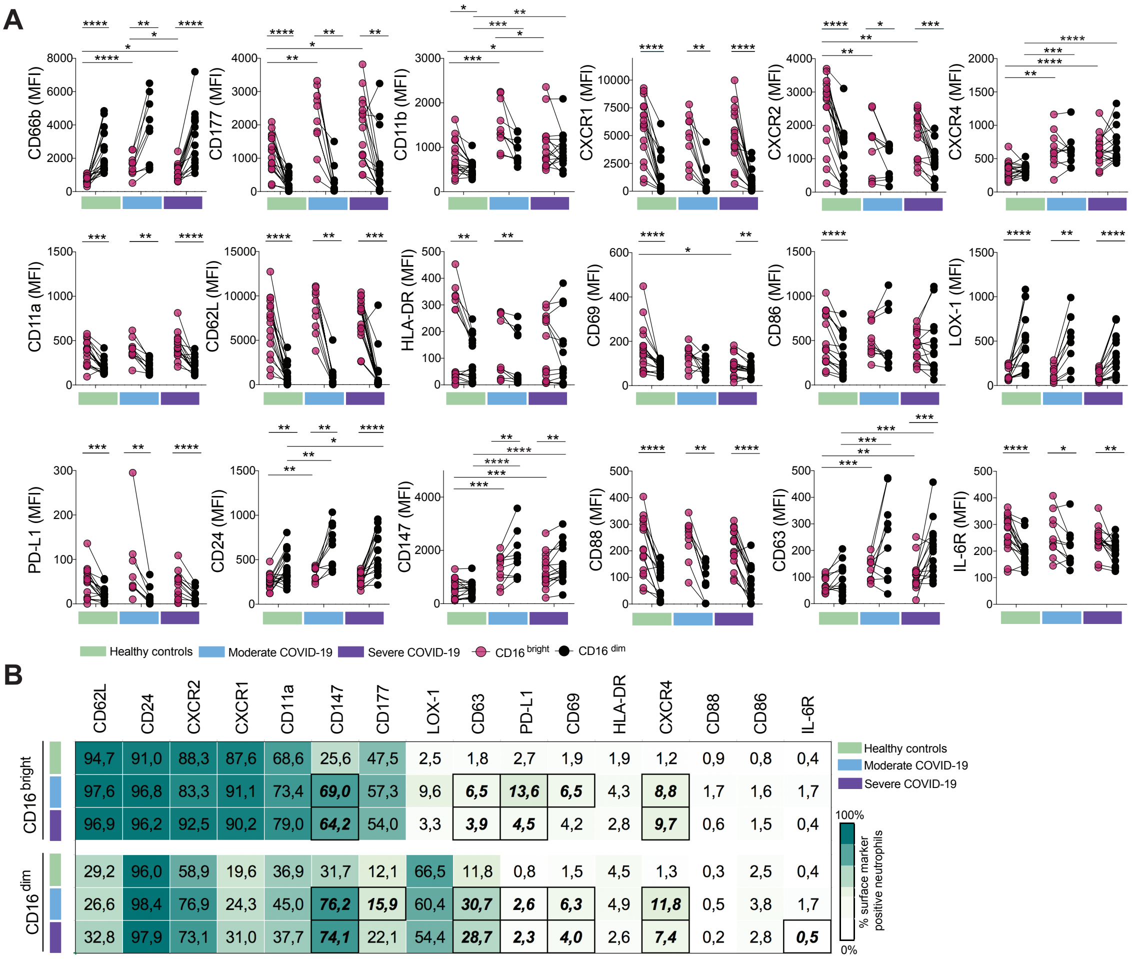

**Supplemental Figure 2**. **Neutrophil phenotypic characterization in COVID-19 patients.** (**A**) Summary scatter plots showing the expression of the indicated markers on CD16$^{bright}$ and CD16$^{dim}$ neutrophils in healthy controls (n=17) moderate COVID-19 patients (n=10) and severe COVID-19 patients (n=16). (**B**) Heatmap showing the fraction within CD16$^{bright}$ and CD16$^{dim}$ neutrophils positive for indicated markers in the three study groups. Mean values (%) are shown and indicated in bold italic if significantly different compared to healthy controls. In (**A**) Wilcoxon matched-pairs signed rank test was used to compare the expression in CD16$^{bright}$ and CD16$^{dim}$ subset within each patient and Kruskall-Wallis test with two-stage Benjamini, Krieger and Yekutieli test was used to compare marker expression across different experimental groups. In (**B**) Kruskall-Wallis test with two-stage Benjamini, Krieger and Yekutieli test was used. FDR adjusted p-values are indicated**.** * $p < 0.05$; ** $p < 0.01$; *** $p < 0.001$; **** $p < 0.0001$.

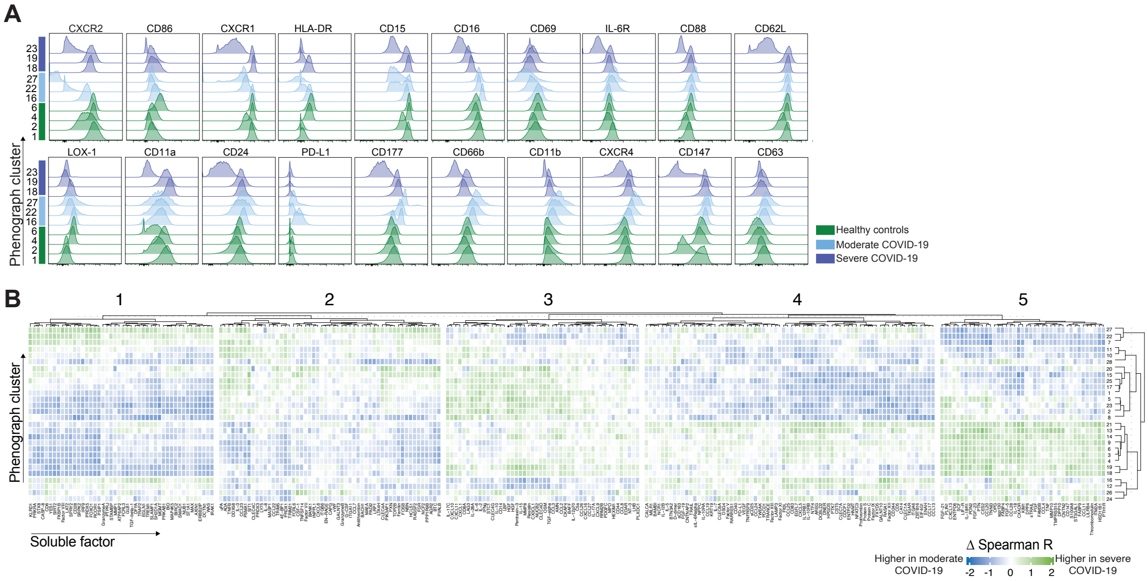

**Supplemental Figure 3**. **Phenograph analysis highlights phenotypic neutrophil heterogeneity and distinct association patterns with soluble factors in moderate and severe COVID-19 patients.** (**A**) Representative histograms showing indicated marker expression for phenograph clusters shown in Figure 3B. (**B**) Heatmap showing the residuals of Spearman r correlation values between phenograph cluster frequencies in each individual and the serum/plasma level of the indicated soluble factors observed in severe minus that observed in moderate COVID-19 patients. The derived residual value (Δ Spearman R) ranges from -2 to +2, and indicates a higher correlation observed in either moderate (when closer to -2) or severe (when closer to +2) COVID-19 of the indicated cluster/soluble factor pairs. To highlight sets of specific associations between cluster frequencies and soluble factors in moderate versus severe COVID-19 patients, the matrix was split by k-means clustering. Raw Δ Spearman r values shown in (**B**) and are provided in detail in Supplemental Table 5.

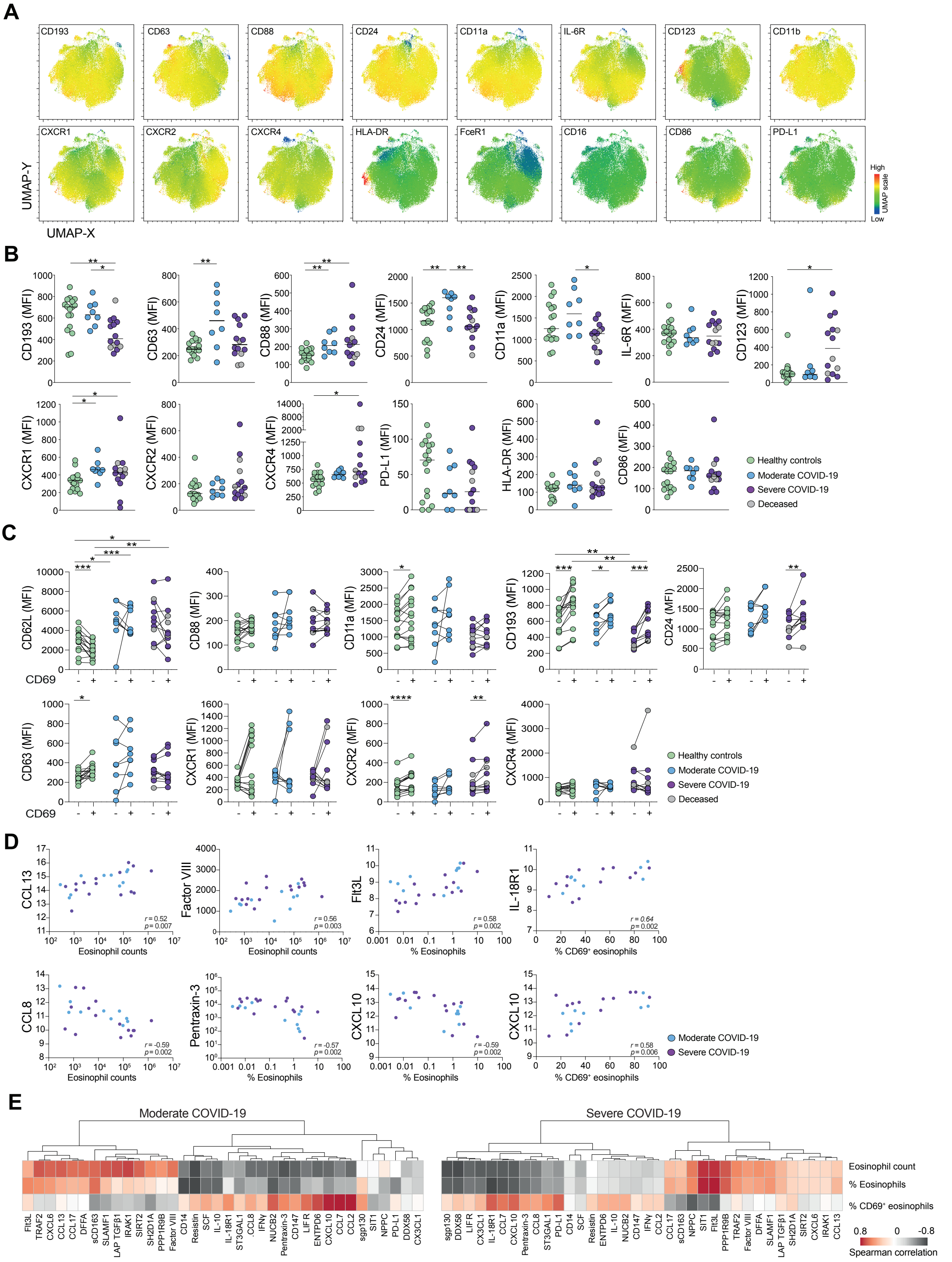

**Supplemental Figure 4**. **Eosinophil phenotype and correlation with soluble factors in moderate and severe COVID-19 patients.** (**A**) Overall UMAP displaying the MFI of the indicated receptors on eosinophils. (**B**) Scatter plots showing the MFI of eosinophil receptors in healthy controls (n=17) and in patients with moderate (n=8) and severe (n=14) COVID-19. The median value for each group is indicated. The deceased patients are indicated in gray. (**C**) Scatter plots showing the MFI of selected receptors in CD69$^-$ and CD69$^+$ eosinophil subsets in moderate (n=8) and severe (n=11) COVID-19 patients compared to healthy controls (n=15). (**D**) Selected scatter plots of representative correlations of eosinophil absolute counts or frequencies with measured soluble factors. (**E**) Heatmaps demonstrating significant correlations ($r < -0.4$ or $r > 0.4$, $p < 0.05$) between absolute eosinophil counts/percent of eosinophils in total leukocytes/percent of $CD69^+$ eosinophils and selected soluble factors in patients with moderate or severe COVID-19. Significant differences between healthy controls and patient groups in (**B**) and (**C**) were evaluated with Kruskall-Wallis test and two-stage Benjamini, Krieger and Yekutieli test. (**C**) Significant differences between paired $CD69^-$ and $CD69^+$ subsets in the same sample were evaluated with Wilcoxon matched-pairs signed rank test. Significant correlations in (**D-E**) were evaluated with Spearman non-parametric test. FDR adjusted p-values are indicated**.** * $p < 0.05$; ** $p < 0.01$; *** $p < 0.001$.

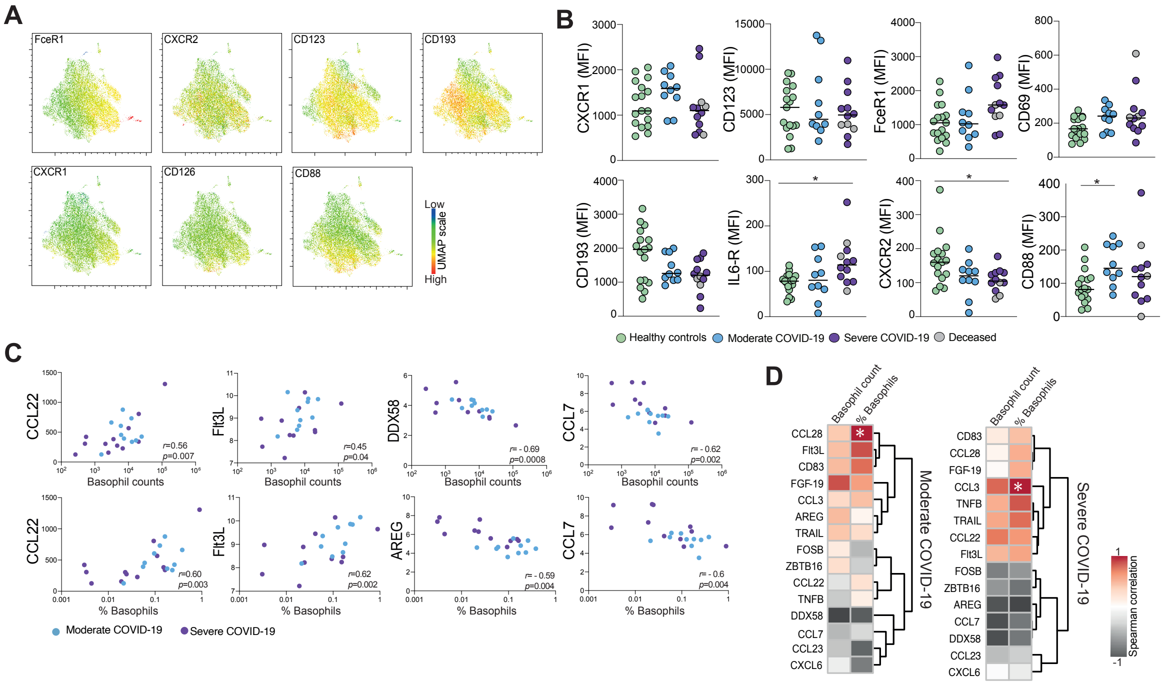

**Supplemental Figure 5**. **Basophil phenotype and relation to circulating soluble factors in COVID-19 patients**. (**A**) Basophil UMAP showing the expression intensity of the indicated markers. (**B**) MFI for selected markers in basophil cell population in heathy controls (n=17), moderate (n=10) and severe COVID-19 patients (n=12). Medians are indicated. (**C**) Selected scatter plots of representative correlations of basophil absolute counts/frequencies with measured soluble factors in moderate and severe COVID-19. (**D**) Heatmaps showing correlation between absolute basophil counts/frequencies and soluble factors, in moderate and severe COVID-19. Significant differences between healthy controls and patient groups in (**B**) were evaluated with Kruskall-Wallis test and two-stage Benjamini, Krieger and Yekutieli test. Significant correlations in (**C-D**) were evaluated with Spearman non-parametric test. FDR adjusted p-values are indicated**.** * $p < 0.05$.

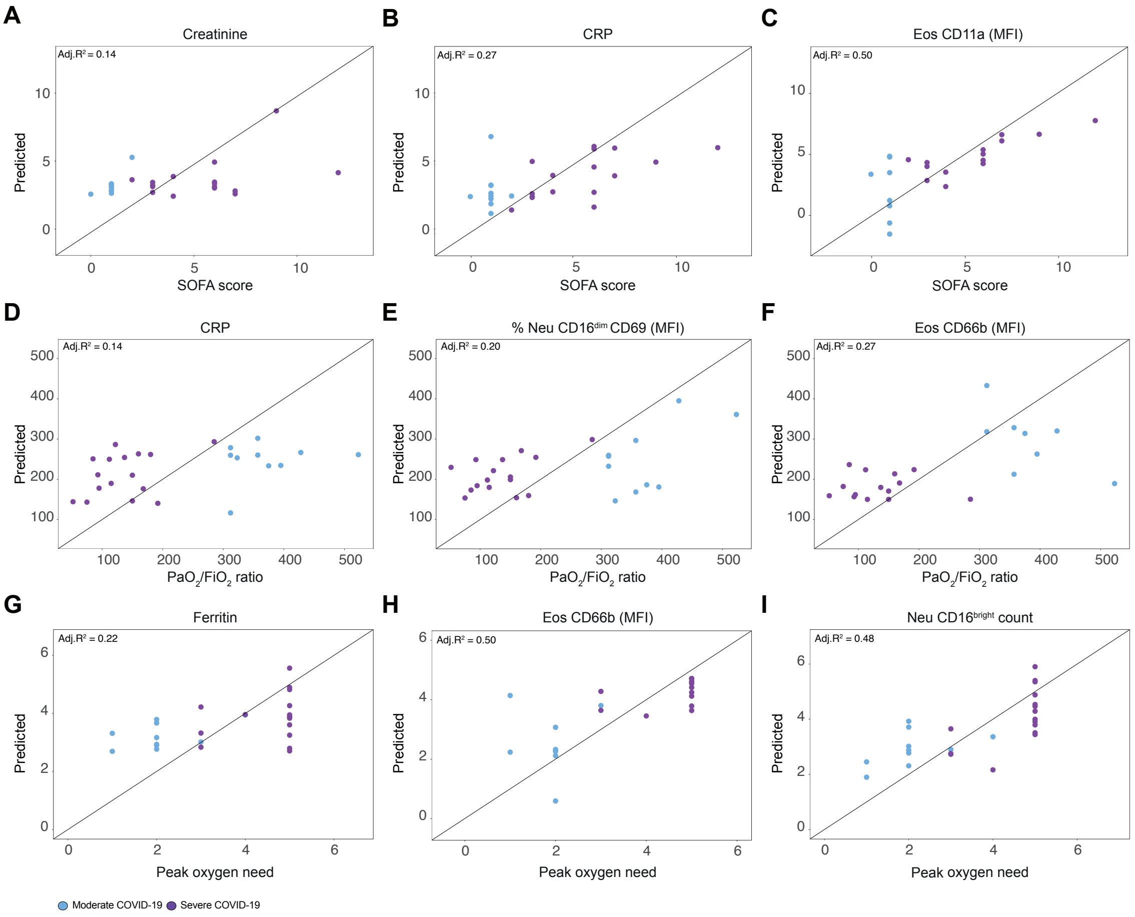

**Supplemental Figure 6. Neither immunologic nor clinical measurements alone are sufficient to predict relevant clinical outcomes**. (**A-I**) Scatter plots of actual values versus predicted values of the linear models for prediction of SOFA score (**A-C**), $PaO_2/FiO_2$ ratio (**D-F**) and peak oxygen need (**G-I**) were modeled based on individual variables. Adj $R^2$, Adjusted $R^2$; Eos, Eosinophils; Neu, Neutrophils.

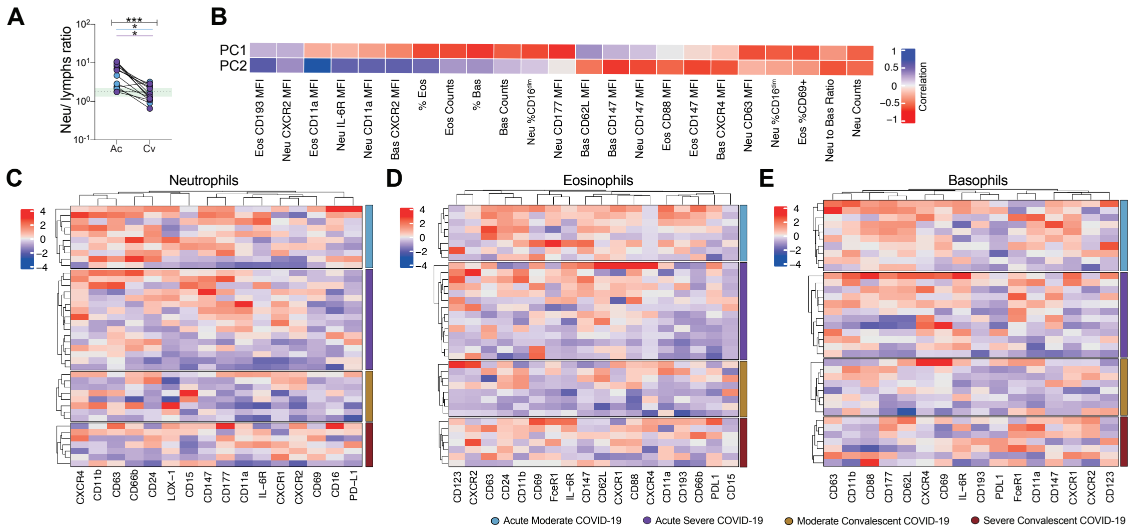

**Supplemental Figure 7. Granulocyte phenotype in paired acute and convalescent samples from COVID-19 patients**. (**A**) Neutrophil (Neu) to lymphocyte (lymphs) ratio, based on absolute cell counts, in paired acute (Ac) and convalescent (Cv) patient samples (n=15). Healthy controls (n=28, median±IQR) ranges are shown in green. Bars indicate statistical significance considering all sampled patients (black), only moderate (blue) or only severe (purple) patients. (**B**) Contribution of individual variables, based on correlation coefficient, to PC1 and PC2 for PCA shown in Figure 7B. (**C**-**E**) Heatmaps showing normalized expression (*z*-score) of all markers in acute moderate/severe and convalescent moderate/severe COVID-19 patients, for neutrophils (**C**), eosinophils (**D**) and basophils (**E**) respectively. In (**A**), Wilcoxon matched-pairs rank test; * $p < 0.05$; ** $p < 0.01$; *** $p < 0.001$; **** $p < 0.0001$.

**Supplemental Table 1.** Inclusion and exclusion criteria of COVID-19 patients and healthy controls.

| | | **Healthy controls n=17** | **Moderate COVID-19 n=10** | **Severe COVID-19 n=16** |
|---|---|---|---|---|
| **INCLUSION** | **Age** | 18-78 | 18-78 | 18-78 |
| | **Male** | 12 (71%) | 7 (70%) | 13 (81%) |
| | **SARS-CoV-2 PCR** | Not assessed | Positive, from nasopharynx or sputum | Positive, from nasopharynx or sputum |
| | **Disease status** | Subjective health, no ongoing symptoms | Ongoing **hospital care** due to COVID-19 disease | Ongoing **intensive care** due to COVID-19 disease |
| | **Days from symptom onset to admission** | | 5-24 days | 5-24 days |
| | **Days from hospital admission to sampling** | | 0-8 days | 0-8 days * |
| | **Respiratory function** | Not assessed | Blood oxygen saturation 90-94% at screening. If supplemental oxygen flow, maximum: 3 L/min | Ongoing mechanical ventilation |
| | **Pre-existing health conditions** | Not assessed | No ongoing malignancy | No ongoing malignancy |
| | **Ongoing medications** | Not assessed | Low level corticosteroids and new drugs aimed specifically at COVID-19 were allowed, but no other immunosuppressant drugs. | Low level corticosteroids and new drugs aimed specifically at COVID-19 were allowed, but no other immunosuppressant drugs. |
| **EXCLUSION** | **SARS-CoV-2 serology** | Positive | | |

* one case was sampled at later time point, 20 days after admission

**Supplemental Table 2.** Clinical characteristics of COVID-19 patients[1].

| **Clinical parameters** | **Moderate COVID-19, n=10** | **Severe COVID-19, n=16** [#] | **p-value*** |
|---|---|---|---|
| **Risk factors** | | | |
| Age | 56 (18-76) | 58 (40-78) | 0.49 |
| Male (sex) | 7 (70%) | 13 (81%) | 0.64 |
| Smoking | 3 (60%) (n=5) | 7 (47%) (n=15) | >0.99 |
| Body mass index | 27.6 (23-35) | 29 (23-55) | 0.33 |
| **Co-morbidity** | | | |
| None | 4 (40%) | 5 (31%) | 0.69 |
| Type 2 diabetes mellitus | 3 (30%) | 4 (25%) | >0.99 |
| Hypertensive heart disease | 2 (20%) | 4 (25%) | >0.99 |
| Coronary heart disease | 2 (20%) | 2 (13%) | 0.63 |
| Asthma | 1 (10%) | 2 (13%) | >0.99 |
| Obesity | 0 | 2 (13%) | 0.51 |
| Obstructive sleep apnea | 1 (10%) | 1 (6%) | >0.99 |
| Chronic pain | 1 (10%) | 1 (6%) | >0.99 |
| **Days from symptom to admission** | 8.5 (4-14) | 9 (3-14) | 0.95 |
| **Days from symptom to sampling** | 13.5 (6-17) | 14.5 (5-24) | 0.24 |
| **Viremia at sampling** | 4 (40%) | 7 (44%) | >0.99 |
| **Serology at sampling** | | | |
| Positive (>=12 AU/mL) | 5 (50%) | 13 (81%) | |
| Borderline (8-11.99 AU/mL) | 2 (20%) | 2 (20%) | |
| Negative (<8 AU/mL) | 3 (30%) | 1 (6%) | |
| SARS-CoV-2 IgG (AU/mL) | 19 (0.26-106) | 55 (3.4-211) | 0.17 |
| **Symptoms at admission** | | | |
| Fever | 10 (100%) | 16 (100%) | >0.99 |
| Cough | 9 (90%) | 13 (81%) | >0.99 |
| Dyspnea | 10 (100%) | 16 (100%) | >0.99 |
| Myalgia | 3 (30%) | 8 (50%) | 0.43 |
| Gastrointestinal symptoms | 1 (10%) | 3 (19%) | >0.99 |
| Embolism/thrombosis | 0 (0%) | 3 (19%) | 0.27 |
| **Positive blood culture** | 0 (0%) | 3 (19%)[†] | 0.27 |

| | | | |
|---|---|---|---|
| **Positive lower respiratory tract culture** | 0 (0%) | 6 (38%)‡ | 0.05 |
| **Peak supportive oxygen therapy** | | | |
| None | 2 (20%) | 0 (0%) | 0.14 |
| Supportive oxygen | 8 (80%) | 4 (25%) | 0.01 |
| Ventilator | 0 (0%) | 11 (69%) | 0.0007 |
| ECMO | 0 (0%) | 1 (6%) | >0.99 |
| $PaO_2/FiO_2$ (mmHg) | 357 (312-523) | 130 (52-285) | <0.0001 |
| **SOFA score** | 1 (0-1) | 6 (3-12) | <0.0001 |
| **Treatment prior to sampling** | | | |
| Anticoagulant prophylaxis | Low mw heparin (n=10; 100%) | Low mw heparin (n=14), oral anticoagulants (n=1), total 93% | >0.99 |
| Antibiotics | 3 (30%): cefotaxime | 12 (75%): cefotaxime, piperacillin/tazobactam | 0.04 |
| Corticosteroids | 2 (20%) | 12 (75%) | 0.0001 |
| Off-label drugs | 0 (0%) | 2 (13%): Tocilizumab, Anakinra | >0.99 |
| **Outcome** | | | |
| Alive | 10 (100%) | 12 (75%) | 0.14 |
| Days of supportive oxygen therapy | 2.5 (0-108) | 20 (7-170)§ | 0.001 |

[1] Unless otherwise stated, all parameters are indicated as median value (range/%).
### One of the severe patients was treated in a high dependency unit at sampling with extracorporeal membrane oxygenation (ECMO). The rest of the severe patients (n=15) were treated in the Intensive care unit (ICU).
§ One patient was treated with ECMO for 170 days before being transferred to another hospital for lung transplantation.
* Statistical significance was determined by Mann-Whitney test or Fisher's exact test.
† *Staphylococcus aureus (n=1), Streptococcus milleri (n=1), S. aureus + Enterococcus faecalis (n=1)*
‡ *S. aureus* (n=2, *Candida albicans (n=1)*, *Escherichia coli (n=1)*, S. aureus + *Streptococcus dysgalactiae (n=1), S. aureus + C. albicans + Klebsiella pneumonia + Aspergillus fumigatus (n=1)*

**Abbreviations**:
ECMO, Extracorporeal membrane oxygenation; SOFA, Sequential organ failure assessment; mw, molecular weight.

**Supplemental Table 3.** Laboratory characteristics of COVID-19 patients.

| Laboratory parameters | Reference | Moderate COVID-19 | Severe COVID-19 | p-value* |
|---|---|---|---|---|
| **C-reactive protein (mg/L)** | <3 | 104 (20-394) (n=9) | 204 (37-346) | 0.14 |
| **Procalcitonin (μg/L)** | <0,5 | 0.40 (0.12-886) (n=9) | 0.61 (0.16-10) | 0.40 |
| **Leukocytes (x10$^9$/L)** | 3.5-8.8 | 7.7 (1.5-12.6) (n=7) | 12.5 (4-18.4) | 0.007 |
| **Neutrophils (x10$^9$/L)** | 1.6-5.9 | 5.4 (0.9-9.3) | 10.9 (3.2-16.7) | 0.001 |
| **Lymphocytes (x10$^9$/L)** | 1.1-3.5 | 1.2 (0.3-2.4) | 0.7 (0-3-2.3) | 0.03 |
| **Thrombocytes (x10$^9$/L)** | 165-387 | 324 (131-642) (n=9) | 331 (146-579) | 0.58 |
| **D-dimer (mg/L)** | <0.56 | 0.71 (0.36-1.3) (n=9) | 2.1 (0.44-5.4) | 0.0009 |
| **Fibrinogen (g/L)** | 2-4.2 | n/a | 6.3 (n=14) (3.5-10.2) | n/a |
| **Troponin T (ng/L)** | <15 | 6 (5-43) (n=7) | 17.5 (6-434) | 0.01 |
| **Creatinine (μmol/L)** | <90 | 67.5 (43-168) | 76 (36-326) | 0.34 |
| **Lactate dehydrogenase (μkat/L)** | <3.5 | 5.55 (3.8-13.7) | 10.6 (0.89-23) | 0.003 |
| **Albumin (g/L)** | 36-48 | 27 (21-37) (n=7) | 19.5 (16-29) | 0.05 |
| **Myoglobin (μg/L)** | <73 | 23 (22-37) (n=3) | 369.5 (36-6400) (n=15) | n/a |
| **Interleukin-6 (ng/L)** | <7 | 53 (2-151) (n=10) | 174 (29-8093) | 0.004 |
| **Interleukin-1β (ng/L)** | <5 | <5 (n=1) | <5 (<5-16.8) (n=15) | n/a |
| **Interleukin-10 (ng/L)** | <5 | n/a | 24 (8.8-47.7) (n=15) | n/a |
| **Tumor necrosis factor-α (ng/L)** | <12 | n/a | 15.5 (8-39.5) (n=15) | n/a |

[1] Unless otherwise stated, all parameters are indicated as median value (range).
[2] Laboratory parameters were measured +/- 24h from sampling. Interleukins and tumor necrosis factor were measured +/- 5 days from sampling. If a group contains missing data, the number of patients for which data was obtained is indicated as ***n*** within cell.
*Statistical significance was determined by Mann-Whitney test.

Supplemental Tables 4 and 5 are separately provided as Excel spreadsheets.

**Supplemental Table 6.** Efficacy of prediction models after correction for the indicated variables.

| Clinical parameters | | p-value* |
|---|---|---|
| **SOFA** | Female | 0.316 |
| | Age | 0.062 |
| | BMI | 0.261 |
| | Days from symptom to sampling | 0.606 |
| | Smoking | 0.362 |
| | Days from admission to sampling | 0.298 |
| | Co-morbidities | 0.828 |
| **$PaO_2/FiO_2$ ratio** | Female | 0.700 |
| | Age | 0.275 |
| | BMI | 0.537 |
| | Days from symptom to sampling | 0.605 |
| | Smoking | 0.069 |
| | Days from admission to sampling | 0.805 |
| | Co-morbidities | 0.214 |
| **Peak oxygen** | Female | 0.925 |
| | Age | 0.850 |
| | BMI | 0.383 |
| | Days from symptom to sampling | 0.396 |
| | Smoking | 0.602 |
| | Days from admission to sampling | 0.240 |
| | Co-morbidities | 0.508 |

*Anova test.

**Supplemental Table 7.** Flow cytometry panel used in this study.

| Antigen | Clone | Fluorochrome | Laser line | Filter | Dilution | Company |
|---|---|---|---|---|---|---|
| **CD16** | 3G8 | BUV805 | *UV (355nm, 100mW)* | 810/40 | 25 | BD |
| **CD86** | 2331 (FUN-1) | BUV737 | | 735/30 | 50 | BD |
| **CD62L** | DREG-56 | BUV661 | | 670/25 | 50 | BD |
| **CD147** | HIM6 | BUV615 | | 605/20 | 25 | BD |
| **PD-L1** | MIH1 | BUV563 | | 580/20 | 20 | BD |
| **CD24** | ML5 | BUV496 | | 515/30 | 25 | BD |
| **CD193** | 5E8 | BUV395 | | 379/28 | 50 | BD |
| **CD63** | H5C6 | BV786 | *Violet (405nm, 100mW)* | 810/40 | 25 | BD |
| **CD15** | W6D3 | BV750 | | 750/30 | 20 | BD |
| **CD88** | D53-1473 | BV711 | | 710/50 | 25 | BD |
| **CD11a** | HI111 | BV650 | | 677/20 | 50 | BD |
| **FceR1** | AER-37 | BV605 | | 605/40 | 50 | BD |
| **LIVE/DEAD Dead Cell Stain** | N/A | Fixable yellow | | 586/15 | 400 | ThermoFisher |
| **CD3** | SK7 | BV510 | | 525/50 | 50 | BioLegend |
| **CD14** | M5E2 | | | | 100 | BioLegend |
| **CD19** | SJ25C1 | | | | 50 | BD |
| **CD56** | NCAM16.2 | | | | 100 | BD |
| **CD304** | U21-1283 | | | | 50 | BD |
| **LOX-1** | 15C4 | BV421 | | 450/50 | 25 | BioLegend |
| **CD69** | FN50 | BB700 | *Blue (488nm, 200nW)* | 710/50 | 25 | BD |
| **CD66b** | G10F5 | BB515 | | 530/30 | 200 | BD |
| **CD123** | 7G3 | PE-Cy7 | *Yellow/green (561nm, 200mW)* | 780/60 | 20 | BD |
| **CXCR4** | 2B11 | PE-Cy5.5 | | 710/50 | 25 | BD |
| **CXCR1** | 8F1/CXCR1 | PE-Cy5 | | 670/30 | 20 | BD |
| **CD11b** | ICRF44 | PE-CF594 | | 610/20 | 100 | BD |
| **CD126** | M5 | PE | | 586/15 | 50 | BD |
| **CXCR2** | 5E8/CXCR2 | APC-Fire | *Red (637nm, 140mW)* | 780/60 | 50 | BioLegend |
| **HLA-DR** | G46-6 | APC-R700 | | 730/45 | 100 | BioLegend |
| **CD177** | MEM-166 | Alexa647 | | 670/30 | 100 | BioLegend |

**Supplemental Acknowledgements**

**List of authors included in the Karolinska KI/K COVID-19 Study Group in alphabetical order:**

Mira Akber[1], Soo Aleman[2], Lena Berglin[1], Helena Bergsten[1], Niklas K Björkström[1], Susanna Brighenti[1], Demi Brownlie[1], Marcus Buggert[1], Marta Butrym[1], Benedict J Chambers[1], Puran Chen[1], Martin Cornillet[1], Angelica Cuapio[1], Isabel Diaz Lozano[1], Lena Dillner[2], Majda Dzidic[1], Johanna Emgård[1], Lars I Eriksson[3], Anna Färnert[2], Malin Flodström-Tullberg[1], Hedvig Glans[2], Jean-Baptiste Gorin[1], Sara Gredmark-Russ[1], Jonathan Grip[3], Alvaro Haroun-Izquierdo[1], Elisabeth Henriksson[1], Laura Hertwig[1], Sadaf Kalsum[1], Tobias Kammann[1], Jonas Klingström[1], Efthymia Kokkinou[1], Egle Kvedaraite[1], Hans-Gustaf Ljunggren[1], Marco Giulio Loreti[1], Magdalini Lourda[1], Kimia T Maleki[1], Karl-Johan Malmberg[1], Nicole Marquardt[1], Johan Mårtensson[3], Christopher Maucourant[1], Jakob Michaëlsson[1], Jenny Mjösberg[1], Kirsten Moll[1], Jagadeeswara Rao Muvva[1], Pontus Nauclér[2], Anna Norrby-Teglund[1], Laura M Palma Medina[1], Tiphaine Parrot[1], André Perez-Potti[1], Björn P Persson[3], Lena Radler[1], Emma Ringqvist[1], Olga Rivera-Ballesteros[1], Olav Rooyackers[3], Johan K Sandberg[1], John Tyler Sandberg[1], Takuya Sekine[1], Ebba Sohlberg[1], Tea Soini[1], Anders Sönnerborg[2], Kristoffer Strålin[2], Mattias Svensson[1], Janne Tynell[1], Christian Unge[4], Renata Varnaite[1], Andreas von Kries[1], David Wullimann[1]

[1]Center for Infectious Medicine, Department of Medicine Huddinge, Karolinska Institutet, Karolinska University Hospital, Stockholm, Sweden

[2]Department of Infectious Diseases, Karolinska University Hospital, Stockholm, Sweden

[3]Function Perioperative Medicine and Intensive Care, Karolinska University Hospital, Stockholm, Sweden

[4]Department of Emergency Medicine, Karolinska University Hospital, Stockholm, Sweden
